# Comparative Effectiveness of the Bivalent (Original/Omicron BA.4/BA.5) mRNA COVID-19 Vaccines mRNA-1273.222 and BNT162b2 Bivalent in Adults With Underlying Medical Conditions in the United States

**DOI:** 10.1101/2024.08.06.24311559

**Authors:** Hagit Kopel, Van Hung Nguyen, Alina Bogdanov, Isabelle Winer, Catherine Boileau, Thierry Ducruet, Ni Zeng, Jessamine Winer-Jones, Daina B. Esposito, Mary Bausch-Jurken, Ekkehard Beck, Mac Bonafede, James A. Mansi

## Abstract

This retrospective cohort study evaluated the relative vaccine effectiveness (rVE) of two bivalent (Original/Omicron BA.4/BA.5) vaccines mRNA-1273.222 versus BNT162b2 Bivalent in preventing COVID-19-related outcomes in adults with underlying medical conditions associated with increased risk for severe COVID-19.

In a linked EHR-claims dataset, US adults (≥18 years) with ≥1 underlying medical condition of interest who received either bivalent vaccine between August 31, 2022, and February 28, 2023, were identified. Inverse probability of treatment weighting was used to adjust for cohort differences. Cohorts were followed up for COVID-19-related hospitalizations and outpatient encounters until May 31, 2023. Hazard ratios and rVEs were estimated using Cox regression. Subgroup analyses were performed on individuals with pre-specified comorbid conditions.

757,572 mRNA-1273.222 and 1,204,975 BNT162b2 Bivalent recipients were identified. The adjusted rVE over a median follow-up of 198 days was 10.9% (6.2%-15.2%) against COVID-19-related hospitalization and 3.2% (1.7%-4.7%) against COVID-19-related outpatient encounters. rVE estimates for COVID-19 hospitalizations among subgroups with comorbid conditions were: diabetes 15.1% (8.7%–21.0%), cardiovascular disease 14.7% (9.0%–20.1%), chronic lung disease 11.9% (5.1%–18.2%), immunocompromised 15.0% (7.2%–22.2%), chronic kidney disease 8.4% (0.5%–15.7%).

Overall, among adults with underlying medical conditions, mRNA-1273.222 was more effective than BNT162b2 Bivalent, especially in preventing COVID-19-related hospitalizations.

## Introduction

Since its emergence in late 2019, the SARS-CoV-2 virus has been continuously evolving and diverging from the strain used to develop the original COVID-19 vaccines [1]. The emergence of the Omicron variant and subvariants, with their ∼50 mutations that promote transmission and immune evasion, resulted in reduced vaccine protection and increased disease burden, including higher infection and hospitalization rates [2,3].

With the aim of targeting the predominant circulating variants during the Omicron period, the bivalent mRNA vaccines, which included mRNA from the original (wild type) SARS-CoV-2 strain, together with an Omicron subvariant (BA.4/BA.5), were developed [4,5]. The vaccines were authorized for use in the US by the US Food and Drug Administration (FDA) in August 2022 and recommended by the US Centers for Disease Control and Prevention (CDC) for all adults in September 2022 [6].

Although all individuals are susceptible to severe outcomes following SARS-CoV-2 infection, some groups, such as older adults and select racial and ethnic minorities, were identified early in the pandemic as being at a higher risk of severe outcomes [7,8]. In addition, individuals with specific underlying medical conditions were shown to be at an increased risk for severe outcomes. The list of underlying medical conditions associated with increased risk includes immunocompromised conditions, cardiovascular diseases, diabetes, and obesity, some of which are highly prevalent in the US adult population, including young adults. An analysis conducted on the National Health Interview Survey (NHIS) has shown that more than 50% of the US adult population (>18) has been diagnosed with at least one chronic condition [9]. Notably, the majority of US adult individuals (including young adults) hospitalized with COVID-19 had at least one underlying medical condition before and during the Omicron period [10,11]. Thus, optimizing protection against COVID-19-related outcomes, especially among these high-risk populations, has direct public health implications which can reduce the burden in terms of infections, chronic complications, and severe outcomes.

While the two bivalent (Original/Omicron BA.4/BA.5) mRNA vaccines that were available in the US (mRNA-1273.222, developed by Moderna, and BNT162b2 Bivalent, developed by Pfizer BioNTech) during the 2022-2023 season, used a similar vaccine technology, previous analysis has shown that mRNA-1273.222 was more effective than BNT162b2 Bivalent in adults aged ≥18 years old [12]. In this analysis, we used the same methodology to evaluate the relative vaccine effectiveness (rVE) of mRNA-1273.222 versus BNT162b2 Bivalent vaccine in the prevention of COVID-19-related hospitalization and outpatient encounters in adult patients (≥18 years of age) living in the US with ≥1 underlying medical condition associated with an increased risk of severe outcomes from COVID-19.

## Methods

### Data Source

This study leveraged electronic health record (EHR) data from the Veradigm Network EHR linked to healthcare claims sourced from Komodo Health spanning March 1, 2020, through May 31, 2023. This integrated dataset has been previously characterized and used previously in COVID-19 epidemiology and VE research [10,12–14]. The dataset used in this study contains only de-identified data as per the de-identification standard defined in Section §164.514(a) of the Health Insurance Portability and Accountability Act of 1996 (HIPAA) Privacy Rule. As a noninterventional, retrospective database study using data from a certified HIPAA–compliant de-identified research database, approval by an institutional review board was not required.

The Komodo data contains claims sourced directly from payers as well as other sources, such as revenue cycle management platforms and claims clearinghouses. For the purpose of this analysis, the closed claims analysis leveraged all claims captured during a period of continuous health plan enrollment, regardless of claim source. By contrast, the open claims sensitivity analysis leveraged all claims during the study period regardless of health plan enrollment. When multiple claims appear for the same encounter, the claim from the payer superseded the claims from any other source.

### Study Design and Study Population

This retrospective, observational cohort study was designed, implemented, and reported in accordance with Good Pharmacoepidemiological Practice, applicable local regulations, and the ethical principles laid down in the Declaration of Helsinki.

Individuals ≥18 years of age with ≥1 underlying medical condition associated with an increased risk of severe outcomes from COVID-19 as defined by the CDC [15] who had received either the mRNA-1273.222 (50 mcg) or BNT162b2 Bivalent vaccine (30 mcg) between August 31, 2022, and February 28, 2023, were eligible for inclusion in the study. Underlying medical conditions were identified based on medical encounters in the 365 days before vaccination. Codes used to identify the underlying medical conditions are listed in Supplementary Table 1. Codes used to identify vaccination with bivalent mRNA vaccines are reported in Kopel et al. [12]. The index date was defined as the date of receipt of the bivalent vaccine, and the cohort entry date (CED) was defined as 7 days after the index date (Figure 1). The follow-up period for outcomes of interest for each individual participant began on the CED and continued until the first occurrence of an event of interest, disenrollment from medical/ pharmacy plan, receipt of an additional COVID-19 vaccination, or the end of available data (May 31, 2023), whichever occurred first.

**Figure 1.**
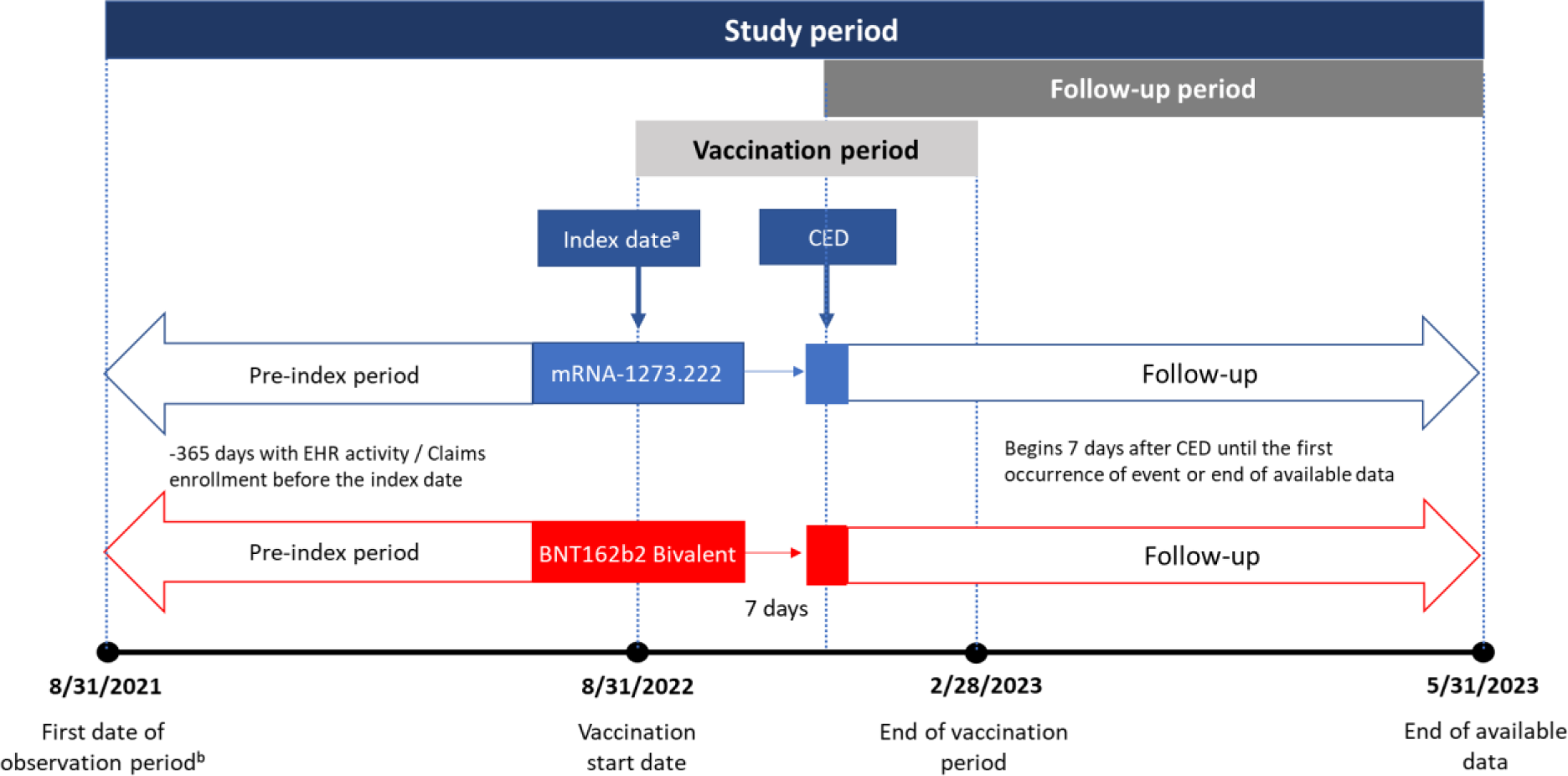
Study Design. CED, cohort entry date. ^a^ The index date is the vaccination date. ^b^ Begins 365 days before vaccination with a bivalent vaccine

Individuals were included in the main analysis if they had a minimum of 365 days of continuous medical and pharmacy claims enrollment in a contributing health plan and ≥1 contact with a health service provider in the previous 365 days. Patients with evidence of COVID-19 infection or additional vaccination between the index date and CED, <1 day of follow-up available, or missing birth year/sex were excluded. The two cohorts were mutually exclusive, and the assignment was based on the first vaccine received during the vaccination period.

### Objectives

The primary objective of this study was to assess the rVE of mRNA-1273.222 versus BNT162b2 Bivalent in preventing COVID-19-related hospitalization. The secondary objective was to assess the rVE of mRNA-1273.222 versus BNT162b2 Bivalent in preventing COVID-19-related outpatient encounters. Both primary and secondary objectives included individuals with any CDC-defined underlying medical condition associated with an increased risk of severe outcomes from COVID-19.

The exploratory objective of this study was to assess the primary and secondary objectives (rVE of COVID-19-related hospitalizations and outpatient encounters) among 5 pre-specified subgroups of patients with underlying medical conditions: cardiovascular disease, chronic kidney disease, chronic lung disease, diabetes, and immunocompromised. These five groups were selected based on their prevalence among the adult population and the risk for severe outcomes [10,15,16]. The specific underlying medical conditions included in these subgroups are defined in Table 1.

**Table 1.**
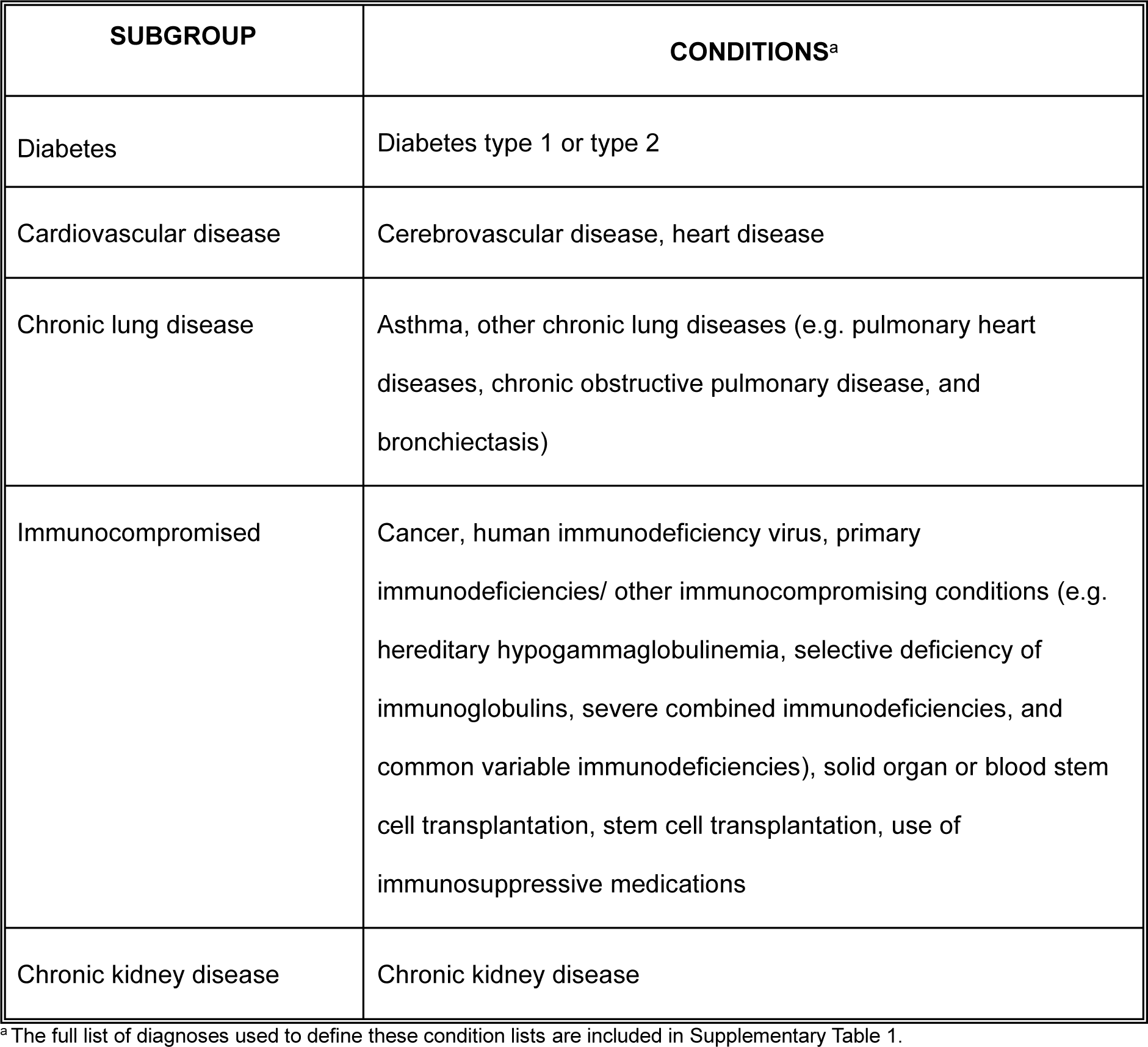
Underlying medical conditions included in the subgroup analyses.

| **SUBGROUP** | **CONDITIONS<sup>a</sup>** |
| --- | --- |
| Diabetes | Diabetes type 1 or type 2 |
| Cardiovascular disease | Cerebrovascular disease, heart disease |
| Chronic lung disease | Asthma, other chronic lung diseases (e.g. pulmonary heart diseases, chronic obstructive pulmonary disease, and bronchiectasis) |
| Immunocompromised | Cancer, human immunodeficiency virus, primary immunodeficiencies/ other immunocompromising conditions (e.g. hereditary hypogammaglobulinemia, selective deficiency of immunoglobulins, severe combined immunodeficiencies, and common variable immunodeficiencies), solid organ or blood stem cell transplantation, stem cell transplantation, use of immunosuppressive medications |
| Chronic kidney disease | Chronic kidney disease |

<sup>a</sup> The full list of diagnoses used to define these condition lists are included in Supplementary Table 1.

### Outcome Measures

The primary outcome of interest was COVID-19-related hospitalizations, which were identified from hospitalization claims with documentation of COVID-19 diagnosis in any position. The secondary outcome of interest was COVID-19-related outpatient encounters, which were identified from outpatient EHR and medical claims with a COVID-19 diagnosis. Outpatient encounters included visits in various clinical settings, such as emergency department visits, urgent care visits, office visits, and telemedicine visits. The codes used to identify outcomes are reported in Kopel et al. [12].

### Covariates

The study utilized patient characteristics as covariates to adjust for any baseline differences between vaccine cohorts (see statistical analysis). The following demographic variables were captured at the index date: age, sex, race, ethnicity, insurance type, region, and month of index. COVID-19 vaccination and infection history were assessed using a lookback period that started on March 1, 2020, and included primary series vaccination, time since the last COVID-19 monovalent vaccination in months, time since the last COVID-19 infection in months, and data source of recorded vaccination. Healthcare resource utilization variables were assessed during the 365-day baseline period and included the number of prior hospitalizations and the number of prior outpatient encounters.

The following underlying medical conditions were captured in the 365 days prior to, and inclusive of, the index date: asthma, cancer, cerebrovascular disease, chronic kidney disease, chronic liver disease, chronic lung disease, cystic fibrosis, dementia, diabetes mellitus, disability, heart conditions, human immunodeficiency virus (HIV), mental health conditions, obesity (body mass index > 30), physical inactivity, primary immunodeficiencies, respiratory tuberculosis, smoking, solid organ or stem cell transplant, and use of select immunosuppressive medications. Pregnancy was captured in the 301 days following the index date. The codes used to identify patients with underlying medical conditions are listed in Supplementary Table 1.

### Statistical Analysis

Statistical analysis followed the methodology described in Kopel et al. [12]. Briefly, propensity scores predicting receipt of mRNA-1273.222 or BNT162b2 Bivalent were calculated using a multivariable logistic model, adjusted for all covariates outlined above. Stabilized and truncated weights were used to re-weight the study sample using the inverse probability of treatment weighting (IPTW) methodology. Standardized mean differences (SMD) were calculated to assess sample balance before and after IPTW. SMDs with absolute values >0.1 indicated covariate imbalance.

Covariates were reported descriptively before and after weighting. Mean and standard deviation (SD) are reported for continuous variables, while number (N) and percent are reported for categorical variables. Follow-up time in days is captured and reported as median and interquartile range (IQR).

Unadjusted hazard ratios (HRs) were reported for the unweighted sample, and adjusted HRs were reported for the weighted sample. Unadjusted HRs were estimated using Cox regression models with exposure as the only predictor. Adjusted HRs were estimated using a multivariable Cox regression model that included the exposure and any baseline covariates with SMD >0.1 after weighting. The rVEs were calculated as 100 × (1 − HR) for both unadjusted and adjusted estimates with 95% confidence intervals (95% CI).

All analyses were performed in the overall population of adults ≥18 years of age with ≥1 underlying medical condition to address the primary objective and secondary objective and then repeated separately for each of the prespecified five cohorts with specific underlying medical conditions of interest (cardiovascular disease, chronic kidney disease, chronic lung disease, diabetes, and immunocompromised).

All statistical analyses were performed using SAS 9.4 or R Statistical Software (v4.1.3) survival (v3.2-13) package.

### Sensitivity analyses

The main analysis was restricted to patients with more than 365 days of continuous health plan enrollment with medical and pharmacy benefits. As this approach may be biased towards patients with stable health insurance, we conducted a sensitivity analysis of the primary and secondary objectives using an open claims approach. The methodology used was the same as the main analysis, except there were no requirements for continuous health plan enrollment. Instead, we required that individuals have at least one medical or pharmacy claim in the 365 days preceding the index date.

A second sensitivity analysis examined the primary and secondary objectives in a shorter follow-up period ending on February 28, 2023. Apart from the shorter follow-up period, all analytical methods were consistent with the main analysis.

## Results

We identified 1,962,547 adults ≥18 years old with at least one underlying medical condition who received a bivalent mRNA vaccine between August 31, 2022, and February 28, 2023, and met all other study criteria (Figure 2). Of these, 757,572 (38.6%) received the mRNA-1273.222 vaccine, and 1,204,975 (61.4%) received the BNT162b2 Bivalent vaccine. Before weighting, the mean (SD) age was 62 (16) years in the mRNA-1273.222 cohort and 60 (16) years in the BNT162b2 Bivalent cohort, and 56.9% and 57.9% of the cohort were female, respectively (Table 2). The most common underlying medical conditions in both cohorts were obesity (mRNA-1273.222: 33.7%; BNT162b2 Bivalent: 34.1%), diabetes (mRNA-1273.222: 33.3%; BNT162b2 Bivalent: 32.2%), and mental health disorders (mRNA-1273.222: 26.7%; BNT162b2 Bivalent: 28.4%). The median (IQR) duration of follow-up was 197 days (147– 225 days) for the mRNA-1273.222 cohort and 200 (148–228) for the BNT162b2 Bivalent cohort.

**Figure 2.**
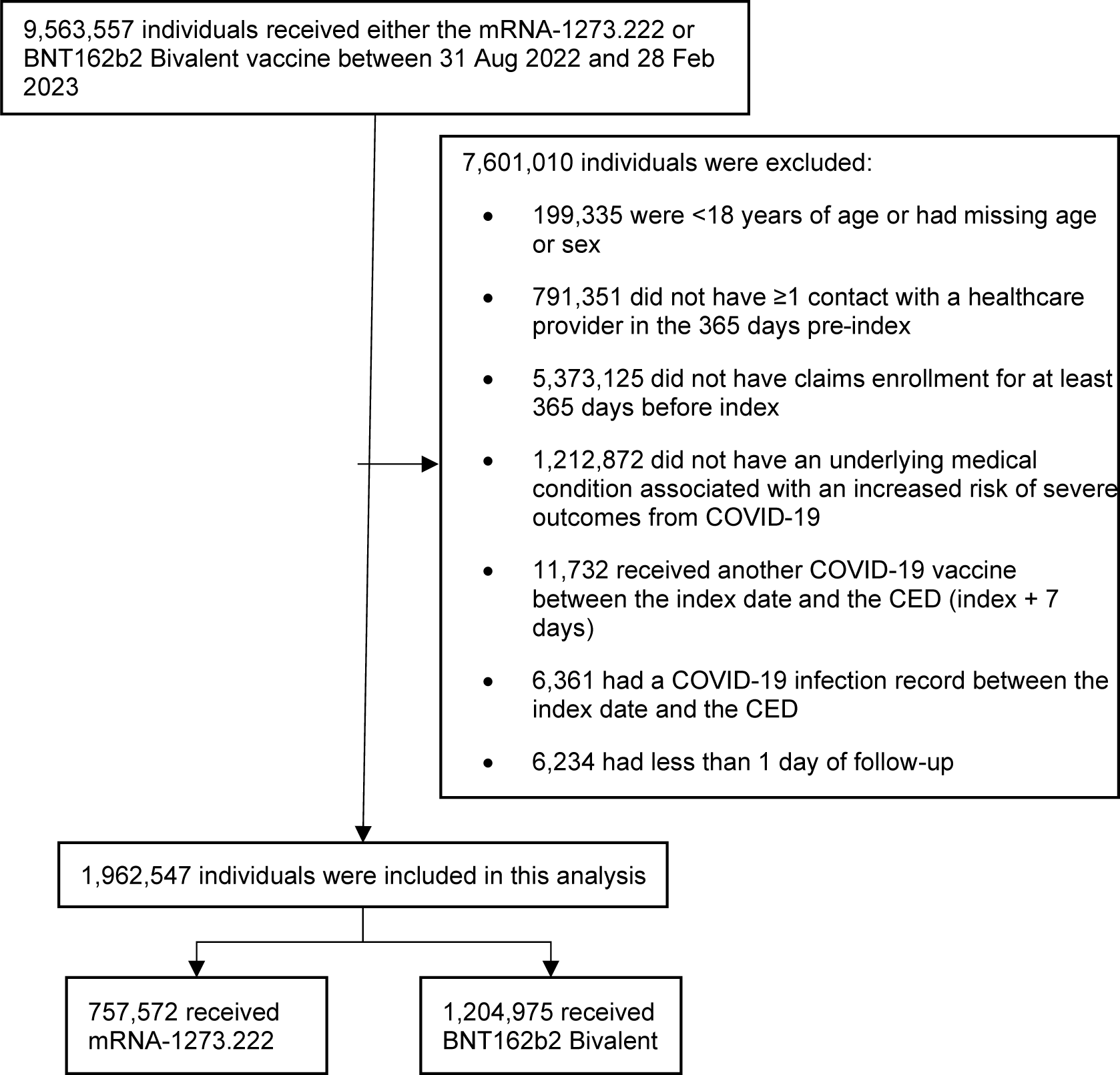
Selection of participants for inclusion in the study.

**Table 2.** Key baseline characteristics of adults with ≥1 underlying medical condition included in the mRNA-1273.222 and BNT162b2 Bivalent vaccine cohorts (pre- and post-weighting).

|  |  | Pre-weighting |  |  | Post-weighting |  |  |
| --- | --- | --- | --- | --- | --- | --- | --- |
|  |  | mRNA-1273.222 | BNT162b2 Bivalent | SMD | mRNA-1273.222 | BNT162b2 Bivalent | SMD |
| **Number of patients** |  | 757,572 | 1,204,975 |  | 758,803 | 1,202,211 |  |
| **Age at index in years, mean (SD)** |  | 62 (16) | 60 (16) | 0.096 | 61 (16) | 61 (16) | 0.003 |
| **Sex, N (%)** | Female | 431,238 (56.9) | 697,587 (57.9) | 0.020 | 436,546 (57.5) | 691,694 (57.5) | <.001 |
|  | Male | 326,334 (43.1) | 507,388 (42.1) |  | 322,257 (42.5) | 510,518 (42.5) |  |
| **Race, N (%)** | Black | 47,522 (6.3) | 82,125 (6.8) | 0.031 | 49,881 (6.6) | 79,383 (6.6) | 0.001 |
|  | Other | 39,457 (5.2) | 60,368 (5.0) |  | 38,699 (5.1) | 61,213 (5.1) |  |
|  | White | 379,026 (50.0) | 589,334 (48.9) |  | 374,392 (49.3) | 592,955 (49.3) |  |
|  | Unknown | 291,567 (38.5) | 473,148 (39.3) |  | 295,831 (39.0) | 468,661 (39.0) |  |
| **Ethnicity, N (%)** | Hispanic | 37,723 (5.0) | 63,334 (5.3) | 0.018 | 38,924 (5.1) | 61,883 (5.1) | <.001 |
|  | Non-Hispanic | 631,927 (83.4) | 997,153 (82.8) |  | 630,037 (83.0) | 997,945 (83.0) |  |
|  | Unknown | 87,920 (11.6) | 144,483 (12.0) |  | 89,842 (11.8) | 142,383 (11.8) |  |
| **Region, N (%)** | Midwest | 153,932 (20.3) | 287,611 (23.9) | 0.089 | 172,470 (22.7) | 272,229 (22.6) | 0.002 |
|  | Northeast | 194,796 (25.7) | 306,893 (25.5) |  | 193,511 (25.5) | 307,058 (25.5) |  |
|  | South | 229,485 (30.3) | 339,329 (28.2) |  | 218,508 (28.8) | 346,982 (28.9) |  |
|  | West | 151,598 (20.0) | 229,463 (19.0) |  | 147,450 (19.4) | 233,374 (19.4) |  |
|  | Unknown | 27,761 (3.7) | 41,679 (3.5) |  | 26,865 (3.5) | 42,569 (3.5) |  |
| **Month of index, N (%)** | 08-2022 | 7 (<0.1) | 14 (<0.1) | 0.068 | 9 (<0.1) | 13 (<0.1) | 0.001 |
|  | 09-2022 | 165,746 (21.9) | 295,324 (24.5) |  | 178,227 (23.5) | 282,665 (23.5) |  |
|  | 10-2022 | 262,769 (34.7) | 417,242 (34.6) |  | 262,976 (34.7) | 416,425 (34.6) |  |
|  | 11-2022 | 161,844 (21.4) | 241,826 (20.1) |  | 156,093 (20.6) | 247,126 (20.6) |  |
|  | 12-2022 | 102,636 (13.5) | 152,770 (12.7) |  | 98,845 (13.0) | 156,509 (13.0) |  |
|  | 01-2023 | 46,739 (6.2) | 70,112 (5.8) |  | 45,090 (5.9) | 71,559 (6.0) |  |
|  | 02-2023 | 17,831 (2.4) | 27,687 (2.3) |  | 17,563 (2.3) | 27,915 (2.3) |  |
| **Primary series COVID-19 vaccine, N (%)** | Heterologous | 64,763 (8.5) | 133,740 (11.1) | 0.175 | 79,300 (10.5) | 123,200 (10.2) | 0.007 |
|  | Homologous | 226,626 (29.9) | 272,401 (22.6) |  | 190,119 (25.1) | 302,057 (25.1) |  |
|  | Not reported | 466,183 (61.5) | 798,834 (66.3) |  | 489,385 (64.5) | 776,955 (64.6) |  |
| **Time since last COVID-19 monovalent vaccination, N (%)** | ≤90 days | 12,342 (1.6) | 15,007 (1.2) | 0.231 | 10,456 (1.4) | 16,657 (1.4) | 0.002 |
|  | 91–180 days | 158,876 (21.0) | 161,599 (13.4) |  | 122,413 (16.1) | 193,483 (16.1) |  |
|  | >180 days | 443,211 (58.5) | 725,100 (60.2) |  | 452,371 (59.6) | 717,402 (59.7) |  |
|  | Not reported | 143,143 (18.9) | 303,269 (25.2) |  | 173,564 (22.9) | 274,670 (22.8) |  |
| **Time since last COVID-19 infection, N (%)** | ≤120 days | 32,609 (4.3) | 52,761 (4.4) | 0.030 | 33,104 (4.4) | 52,418 (4.4) | <.001 |
|  | 121–180 days | 17,269 (2.3) | 27,424 (2.3) |  | 17,371 (2.3) | 27,457 (2.3) |  |
|  | >180 days | 67,135 (8.9) | 117,056 (9.7) |  | 71,407 (9.4) | 113,132 (9.4) |  |
|  | Not reported | 640,559 (84.6) | 1,007,734 (83.6) |  | 636,921 (83.9) | 1,009,204 (83.9) |  |
| **Underlying medical conditions, N (%)** | Asthma | 107,684 (14.2) | 175,269 (14.5) | 0.009 | 109,615 (14.4) | 173,562 (14.4) | <.001 |
|  | Cancer | 99,704 (13.2) | 151,325 (12.6) | 0.018 | 96,642 (12.7) | 153,428 (12.8) | <.001 |
|  | Cerebrovascular disease | 68,237 (9.0) | 104,581 (8.7) | 0.012 | 66,354 (8.7) | 105,537 (8.8) | 0.001 |
|  | Chronic kidney disease | 106,577 (14.1) | 164,005 (13.6) | 0.013 | 103,660 (13.7) | 165,086 (13.7) | 0.002 |
|  | Chronic liver diseases | 14,318 (1.9) | 22,958 (1.9) | 0.001 | 14,377 (1.9) | 22,830 (1.9) | <.001 |
|  | Chronic lung diseases<sup>a</sup> | 98,839 (13.0) | 150,479 (12.5) | 0.017 | 95,808 (12.6) | 152,296 (12.7) | 0.001 |
|  | Cystic fibrosis | 286 (<0.1) | 422 (<0.1) | 0.001 | 272 (<0.1) | 431 (<0.1) | <.001 |
|  | Diabetes type 1 & 2 | 252,180 (33.3) | 388,331 (32.2) | 0.023 | 246,489 (32.5) | 391,550 (32.6) | 0.002 |
|  | Disabilities | 82,976 (11.0) | 141,091 (11.7) | 0.024 | 87,274 (11.5) | 137,767 (11.5) | 0.001 |
|  | Heart conditions | 169,131 (22.3) | 256,250 (21.3) | 0.026 | 163,349 (21.5) | 259,743 (21.6) | 0.002 |
|  | HIV | 7,327 (1.0) | 11,394 (0.9) | 0.002 | 7,269 (1.0) | 11,491 (1.0) | <.001 |
|  | Mental health disorders | 201,996 (26.7) | 342,635 (28.4) | 0.040 | 211,585 (27.9) | 334,556 (27.8) | 0.001 |
|  | Neurologic conditions | 26,905 (3.6) | 47,214 (3.9) | 0.019 | 28,561 (3.8) | 45,438 (3.8) | <.001 |
|  | Obesity | 255,376 (33.7) | 410,845 (34.1) | 0.008 | 257,461 (33.9) | 408,164 (34.0) | <.001 |
|  | Physical inactivity | 1,200 (0.2) | 1,941 (0.2) | <.001 | 1,223 (0.2) | 1,931 (0.2) | <.001 |
|  | Primary Immunodeficiencies | 15,989 (2.1) | 25,055 (2.1) | 0.002 | 15,724 (2.1) | 25,043 (2.1) | <.001 |
|  | Pregnancy<sup>b</sup> | 5,072 (0.7) | 9,587 (0.8) | 0.015 | 5,741 (0.8) | 9,048 (0.8) | <.001 |
|  | Smoking<sup>c</sup> | 153,460 (20.3) | 244,998 (20.3) | 0.002 | 153,751 (20.3) | 243,993 (20.3) | <.001 |
|  | Solid organ or hematopoietic cell transplantation | 5,078 (0.7) | 8,679 (0.7) | 0.006 | 5,287 (0.7) | 8,429 (0.7) | <.001 |
|  | Tuberculosis | 444 (0.1) | 719 (0.1) | <.001 | 447 (0.1) | 711 (0.1) | <.001 |
|  | Use of immunosuppressants | 63,796 (8.4) | 98,130 (8.1) | 0.010 | 62,513 (8.2) | 99,075 (8.2) | <.001 |

IQR, interquartile range; SD, standard deviation; SMD, standardized mean difference
<sup>a</sup>except for asthma
<sup>b</sup>Includes recent pregnancy
<sup>c</sup>Includes current and former smoker

Before weighting, the subgroup analyses included 640,511 individuals with diabetes, 516,081 with cardiovascular disease, 474,573 with chronic lung disease, 427,652 who were immunocompromised, and 270,582 with chronic kidney disease (Table 3). This comprised 33.3% of the mRNA-1273.222 cohort and 32.2% of the BNT162b2 Bivalent cohort who were included in the diabetes subgroup analysis, 27.1% and 25.8% who were included in the cardiovascular disease subgroup analysis, 24.2% and 24.1% who were included in the lung disease subgroup analysis, 22.3% and 21.4% who were included in the immunocompromised subgroup analysis, and 14.1% and 13.6% who were included in the chronic kidney disease subgroup analysis, respectively. These subgroups were not mutually exclusive, as individuals may have multiple underlying medical conditions. Median follow-up time in the subgroups ranged from 195 to 199 in the mRNA-1273.222 cohort and from 197 to 202 in the BNT162b2 Bivalent cohort.

**Table 3.**
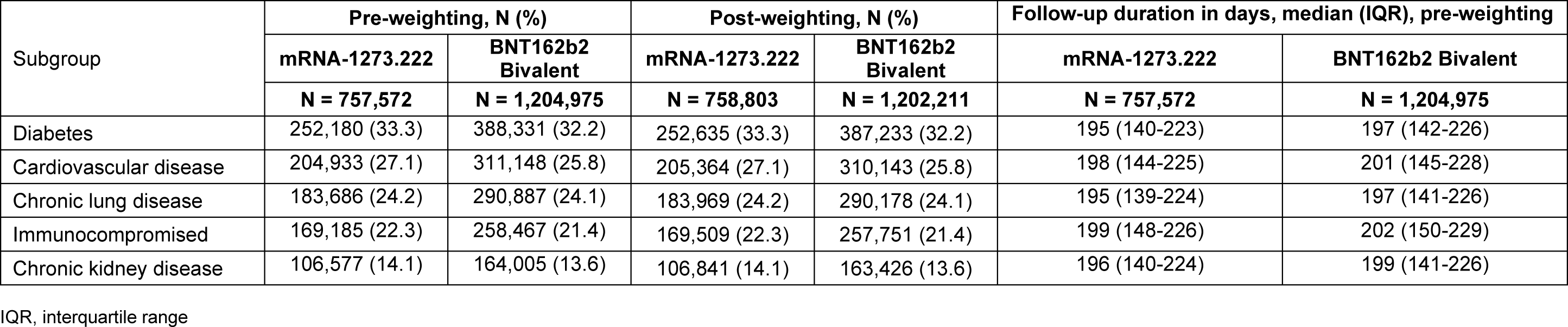
Number of individuals included in the subgroup analyses and duration of follow-up.

Baseline characteristics of the overall analyses and the subgroup analyses before and after weighting are reported in Table 2 and Supplementary Tables 2-6. After weighting, no variables had an SMD > 0.1 in either the overall study cohorts or in the underlying medical condition subgroups.

### Main Analysis

In the overall study cohort, post-weighting, 2,360 (0.31%) individuals who received mRNA-1273.222 and 4,198 (0.35%) who received BNT162b2 Bivalent were hospitalized for COVID-19 during the follow-up period. Whereas, 26,185 (3.5%) individuals who had received mRNA-1273.222 and 42,866 (3.6%) who had received BNT162b2 Bivalent had an outpatient encounter related to COVID-19.

For both the primary and secondary endpoints, mRNA-1273.222 was significantly more effective at preventing the outcomes of interest compared to BNT162b2 Bivalent. Specifically, the rVE (95%CI) against COVID-19-related hospitalization was 10.9% (6.2%– 15.2%, p<0.001), and the rVE against outpatient COVID-19 was 3.2% (1.7%–4.7%, p<0.001) (Figure 3).

**Figure 3.**
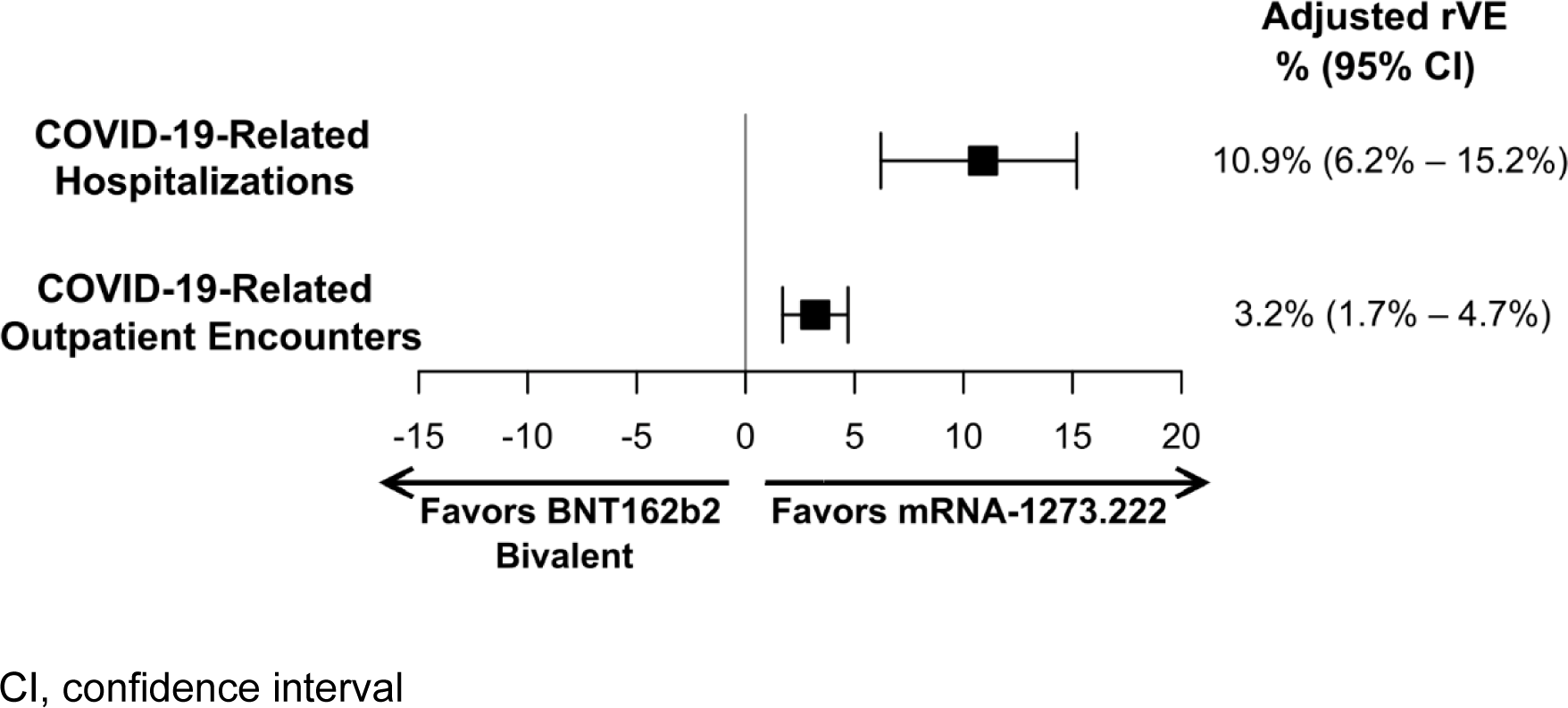
Adjusted rVE estimates of mRNA-1273.222 vs. BNT162b2 Bivalent for adults with ≥1 underlying medical condition.

For all subgroups, mRNA-1273.222 was significantly more effective at preventing COVID-19-related hospitalizations and outpatient encounters compared to BNT162b2 Bivalent (Figure 4). The rVE (95% CI) of mRNA-1273.222 vs. BNT162b2 Bivalent against COVID-19-related hospitalization was 15.1% (8.7%–21.0%) in patients with diabetes, 14.7% (9.0%– 20.1%) in those with cardiovascular disease, 11.9% (5.1%–18.2%) in those with chronic lung disease, 15.0% (7.2%–22.2%) in immunocompromised patients, and 8.4% (0.5%–15.7%) in CKD patients. Point estimates for rVE against outpatient COVID-19 were lower but statistically significant and ranged from 3.1% in immunocompromised patients to 7.6% in patients with CKD. Unadjusted and adjusted hazard ratios and 95% CI are reported in Table 4.

**Figure 4.**
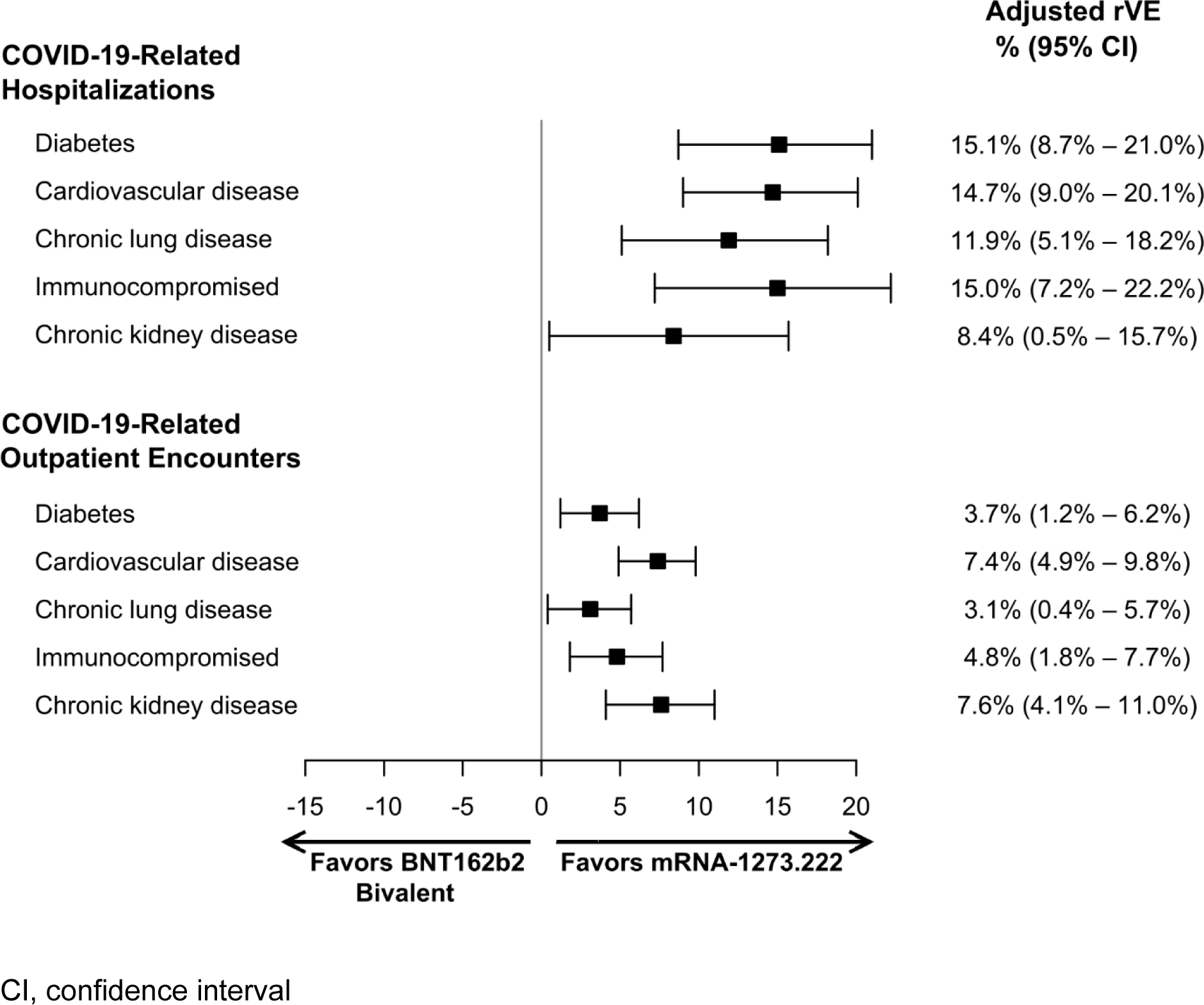
Adjusted rVE estimates of mRNA-1273.222 versus BNT162b2 Bivalent among subgroups with specific underlying medical conditions.

**Table 4.** Unadjusted and adjusted hazard ratios (HR) and 95% confidence intervals (CI) for mRNA-1273.222 versus BNT162b2 Bivalent.

|  | Unadjusted HR<sup>a</sup>, (95% CI) |  | Adjusted HR<sup>b</sup>, (95% CI) |  |
| --- | --- | --- | --- | --- |
|  | COVID-19-related hospitalization | COVID-19-related outpatient encounter | COVID-19-related hospitalization | COVID-19-related outpatient encounter |
| Overall | 0.873 (0.830-0.919) | 0.971 (0.956-0.986) | 0.891 (0.848-0.938) | 0.968 (0.953-0.983) |
| **Subgroup analyses** |  |  |  |  |
| Diabetes | 0.802 (0.746-0.863) | 0.947 (0.923-0.972) | 0.849 (0.790-0.913) | 0.963 (0.938-0.988) |
| Cardiovascular disease | 0.790 (0.739-0.844) | 0.894 (0.871-0.919) | 0.853 (0.799-0.910) | 0.926 (0.902-0.951) |
| Chronic lung disease | 0.852 (0.791-0.918) | 0.957 (0.932-0.984) | 0.881 (0.818-0.949) | 0.969 (0.943-0.996) |
| Immunocompromised | 0.835 (0.764-0.912) | 0.946 (0.917-0.976) | 0.850 (0.778-0.928) | 0.952 (0.923-0.982) |
| Chronic kidney disease | 0.843 (0.775-0.916) | 0.894 (0.860-0.928) | 0.916 (0.843-0.995) | 0.924 (0.890-0.959) |
| **Sensitivity analyses** |  |  |  |  |
| Open claims | 0.859 (0.833-0.887) | 0.928 (0.919-0.937) | 0.880 (0.853-0.907) | 0.869 (0.818-0.922) |
| Closed claims – cut-off 28 Feb 23 | 0.854 (0.804-0.907) | 0.974 (0.957-0.991) | 0.950 (0.941-0.960) | 0.962 (0.946-0.979) |

<sup>a</sup>Pre-weighting <sup>b</sup>post-weighting

### Sensitivity analyses

The results of the sensitivity analysis were consistent with the findings of the main analysis (Tables 4 and 5). Specifically, in the open claims dataset, we identified 1,960,185 adults ≥18 years old with at least one underlying medical condition who received mRNA-1273.222 and 2,969,597 who received BNT162b2 Bivalent vaccine during the vaccination period. Study subjects met all inclusion criteria as in the main analysis except that the requirement for continuous enrollment in claims ≥365 days prior to the index date was replaced with a requirement for at least one claim within 365 days of the index date. In this population, the rVE (95% CI) of mRNA-1273.222 versus BNT162b2 Bivalent against COVID-19-related hospitalization was 12.0% (9.3%–14.7%), and the rVE against outpatient COVID-19 was 5.0% (4.0%–5.9%). Similarly, the use of a shorter follow-up period (ending February 28, 2023) in the closed claims database resulted in rVE estimates of 13.1% (7.8%–18.2%) and 3.8% (2.1%–5.4%) against hospitalized and outpatient COVID-19, respectively. Baseline characteristics of individuals included in the sensitivity analyses before and after weighting are reported in Supplementary Tables 7 and 8.

**Table 5.** Adjusted rVE estimates^a^, sensitivity analyses.

| Outcome | Adjusted rVE |
| --- | --- |
| ***Open claims – cut-off 31 May 23*** |  |
| COVID-19-related hospitalization | 12.0% (9.3% – 14.7%) |
| COVID-19-related outpatient encounter | 5.0% (4.0% – 5.9%) |
| ***Closed claims – cut-off 28 Feb 23*** |  |
| COVID-19-related hospitalization | 13.1% (7.8% – 18.2%) |
| COVID-19-related outpatient encounter | 3.8% (2.1% – 5.4%) |

<sup>a</sup> mRNA-1273.222 versus BNT162b2 Bivalent
rVE, relative vaccine effectiveness

## Discussion

In this real-world retrospective analysis of more than 1.9 million individuals vaccinated with a bivalent COVID-19 vaccine, mRNA-1273.222 was significantly more effective than BNT162b2 Bivalent in preventing COVID-19-related hospitalizations in adults ≥18 years old with at least one underlying medical condition. Similarly, mRNA-1273.222 offered greater protection against outpatient encounters compared to BNT162B2 bivalent but to a lesser extent. These results were consistent in the subgroup analyses of prespecified comorbid conditions, including diabetes, cardiovascular disease, immunocompromised conditions, chronic lung disease, and chronic kidney disease. Notably, the median follow-up duration was greater than 6 months, and results were similar in the sensitivity analysis of shorter follow-up duration. As SARS-CoV-2 continues to evolve, this study highlights the need to reassess vaccine strategies to provide optimal protection for everyone and especially for adults with underlying medical conditions who require comprehensive clinical management and are at higher risk of severe outcomes.

These findings align with our previous analysis of the relative effectiveness rVE of bivalent vaccines in the overall adult population. We reported rVEs (95% CI) of 9.8% (2.6–16.4%) against COVID-19-related hospitalizations and 5.1% (3.2%–6.9%) against COVID-19-related outpatient encounters for mRNA-1273.222 compared to BNT162b2 Bivalent [14]. Among adults aged 65 and older, the estimated rVEs were slightly higher, showing 13.5% (5.5%– 20.8%) against COVID-19-related hospitalizations and 10.7% (8.2%–13.1%) against COVID-19-related outpatient encounters.

Earlier studies assessing the primary series and initial monovalent boosters found similar results, showing greater effectiveness of the mRNA-1273 vaccine over the BNT162b2 vaccine, particularly in groups at higher risk due to age and comorbidities [14,17–19]. These findings have remained consistent across different variant periods for high-risk groups and extend to subpopulations with specific medical conditions. For instance, vaccine effectiveness (VE) against infection and death was higher for the primary series of mRNA-1273 compared to BNT162b2 in a study focusing on individuals with diabetes. Similarly, the effectiveness against medically attended SARS-CoV-2 infection or related hospitalization after a third dose was higher in patients with diabetes, hypertension, heart disease, cancer, or COPD [20,21].

The difference in protection afforded by the vaccines may be a result of a stronger immune response elicited by mRNA-1273 compared to BNT162b2, especially in individuals who are immunocompromised or have comorbid conditions. In a pairwise meta-analysis across various immunocompromising conditions, mRNA-1273 vaccination was more likely to result in seroconversion and higher antibody titers than BNT162b2 vaccination [22]. Additionally, studies have shown a stronger humoral response associated with mRNA-1273 vaccination in type 1 diabetics, dialysis patients, patients with chronic medical conditions, and elderly individuals with multiple comorbid conditions, compared to BNT162b2 vaccination [23–26].

Research has established that individuals with certain underlying medical conditions represent approximately 22% of the adult global population [27]. These individuals are at increased risk of severe outcomes from COVID-19 [28–30], and this risk increases further with the number of conditions [31]. An analysis of a large US hospital all-payer database found that the risk ratio (95% CI) of death among individuals hospitalized with COVID-19 increased from 1.5 (1.4–1.7) for individuals with 1 underlying medical condition to 3.8 (3.5– 4.2) for individuals with more than 10 underlying medical conditions relative to individuals without any underlying medical conditions [31]. As expected, severe COVID-19 outcomes increase the economic burden. Specifically, hospitalization costs for COVID-19 patients with comorbid conditions have been significantly higher when compared to individuals without known underlying conditions. Patients with chronic kidney disease had a 64% higher cost, liver disease patients had a 37% higher cost, and those with cerebrovascular disease had a 30% higher cost compared to those without these conditions [32]. In addition, following the acute phase of COVID-19, patients at higher risk of COVID-19 were also found to have increased overall healthcare spend and cost. A study using US commercial and Medicare Advantage claims of patients at high risk for COVID-19 estimated the mean overall per-patient medical cost during the year following acute COVID-19 at $27,077 which represents an increase of $5,200 or 23.8% in comparison to the medical cost occurring in the year before the COVID-19 infection [33]. Thus, COVID-19 contributes to increasing the already high non-COVID healthcare costs associated with patients at high risk.

In summary, maximizing protection against preventive diseases, especially among patients with underlying medical conditions who require comprehensive clinical management, can reduce the burden of COVID-19 on individuals and the healthcare system.

### Strengths and limitations

The primary strength of this analysis is the depth and breadth of the linked EHR and claims dataset used in this analysis. By integrating sources of patient information across the continuum of care, we increase the likelihood of capturing exposures (vaccination events), covariates (patient characteristics and underlying medical conditions), and outcomes (COVID-19-related hospitalizations and outpatient encounters). Additionally, as information on exposure, outcome, and covariates was collected from patient records in a consistent manner across all cohorts, there is a reduced likelihood of differential misclassification. Residual confounding between cohorts was addressed using propensity score weighting and multivariable regression.

This study is subject to several limitations which are inherent to many non-interventional studies utilizing real-world data. Specifically, this retrospective observational study, which relies on routinely collected data on insured patients, might include data entry errors and cannot be generalized to the uninsured population. Additionally, the current primary analysis was restricted to closed claims, which provide a comprehensive overview of a patient’s healthcare interactions but may not capture all cases and is restricted to patients with stable healthcare insurance. However, the sensitivity analysis performed using an open claims approach was consistent with the main analysis, which supports a broad generalizability of our findings. Furthermore, we required that study subjects have at least one healthcare encounter during the previous 365 days, which would exclude healthy patients; however, as this study was restricted to individuals with underlying medical conditions, the effects of this bias are minimized.

Finally, there may have been differences between the two vaccine groups that were not fully accounted for by the pre-defined covariates and residual confounding may be present between the cohorts. For example, differences in the timing and availability of the two vaccines may have impacted which patients were more likely to receive one vaccine over the other. In our previous analysis that included the overall adult population, we found no difference in the proportion of patients vaccinated with each vaccine over time [12].

## Conclusions

In this real-world analysis, mRNA-1273.222 was significantly more effective than BNT162b2 Bivalent at preventing COVID-19-related hospitalizations and outpatient encounters among adults ≥18 years with at least one underlying medical condition associated with higher risk for COVID-19 severe outcomes.

## Disclosures

### Funding

This work was supported by Moderna Inc. via a contract with Veradigm.

## Conflicts of Interest

H.K., D.B.E., M. B., E. B., and J. M. are employees of and shareholders in Moderna Inc. A.B., J.P.W-J., N.Z., I.H.W., and M.B. are employees of Veradigm, which was contracted by Moderna and received fees for data management and statistical analyses. V.H.N., C.B., and T.D., are employees of VHN Consulting, which was contracted by Moderna to help conduct this analysis.

## Acknowledgments

Christopher Adams, an employee of Veradigm, provided programming support and quality assurance for this analysis.

## Data Availability Statement

The data that support the findings of this study were used under license from Veradigm and Komodo Health. Due to data use agreements and its proprietary nature, restrictions apply regarding the availability of the data. Further information is available from the corresponding author.

## Author Contributions

Conceptualization, J.A.M., H.K., A.B.A., M.B., I.W., and V.H.N.; methodology, all authors; validation, A.B., J.P.W.-J., N.Z., and I.H.W.; formal analysis, A.B., J.P.W.-J., N.Z., and I.H.W.; investigation, A.B., J.P.W.-J., N.Z., and I.H.W.; data curation, N.Z.; writing—original draft preparation, J.P.W.-J., J.A.M., H.K. and V.H.N; writing—review and editing, all authors; visualization, J.P.W.-J., J.A.M., and H.K.; All authors have read and agreed to the published version of the manuscript.

## Ethics approval and consent to participate

The linked dataset only contains de-identified data as per the de-identification standard defined in Section §164.514(a) of the Health Insurance Portability and Accountability Act of 1996 (HIPAA) Privacy Rule. The process by which the data is de-identified is attested to through a formal determination by a qualified expert as defined in Section §164.514(b)(1) of the HIPAA Privacy Rule. Because this study used only de-identified patient records, it is therefore no longer subject to the HIPAA Privacy Rule and is therefore exempt from Institutional Review Board approval and for obtaining informed consent according to US law. This study was conducted in compliance with the Declaration of Helsinki and used only de-identified data.

---

**Supplementary Table 1.**
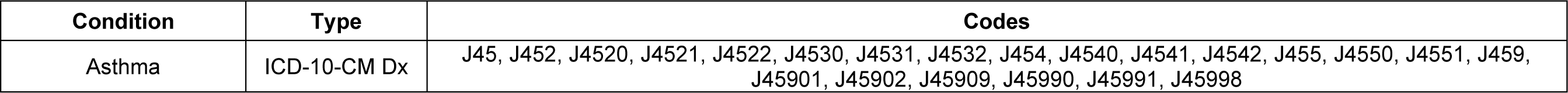
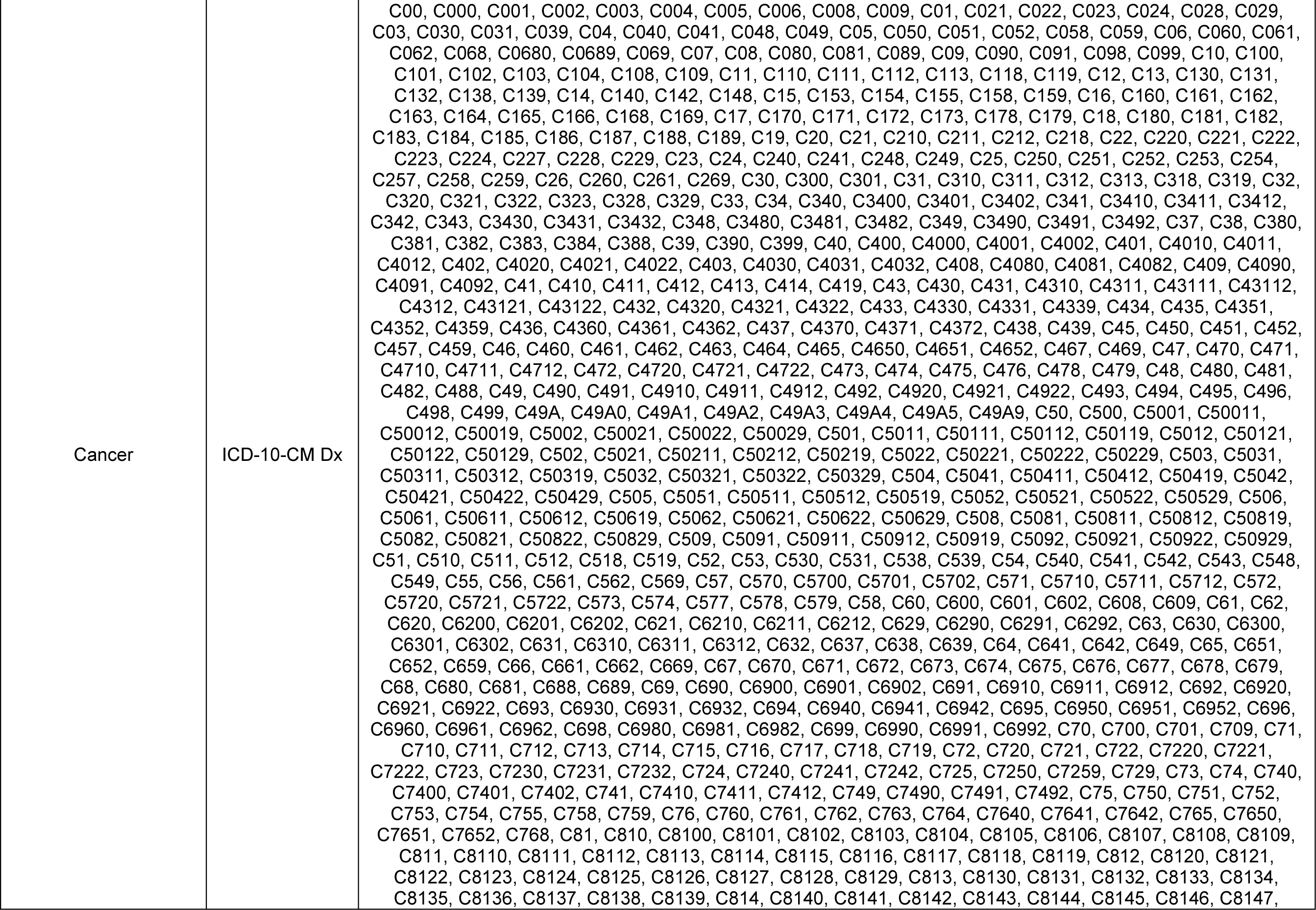
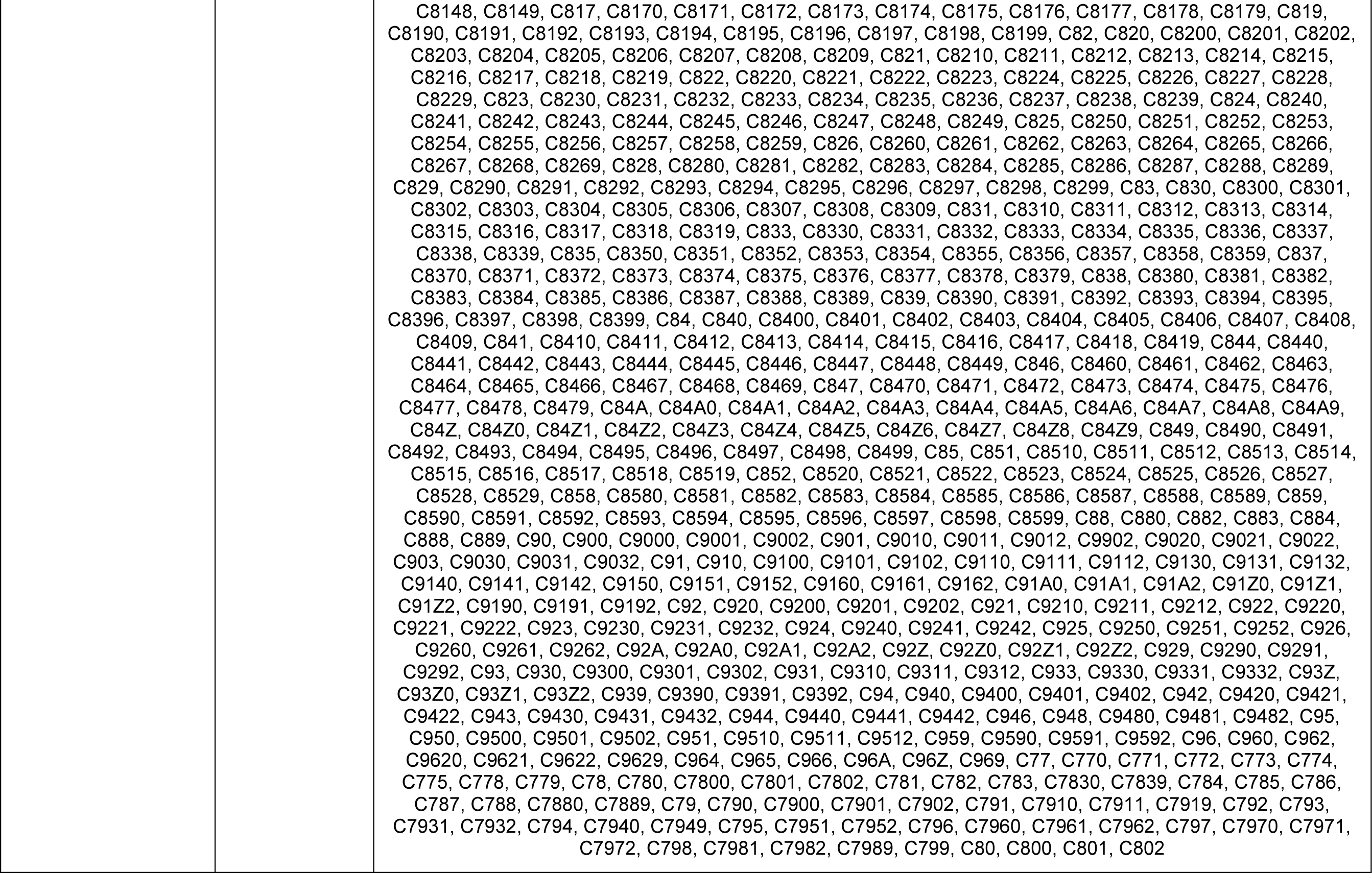
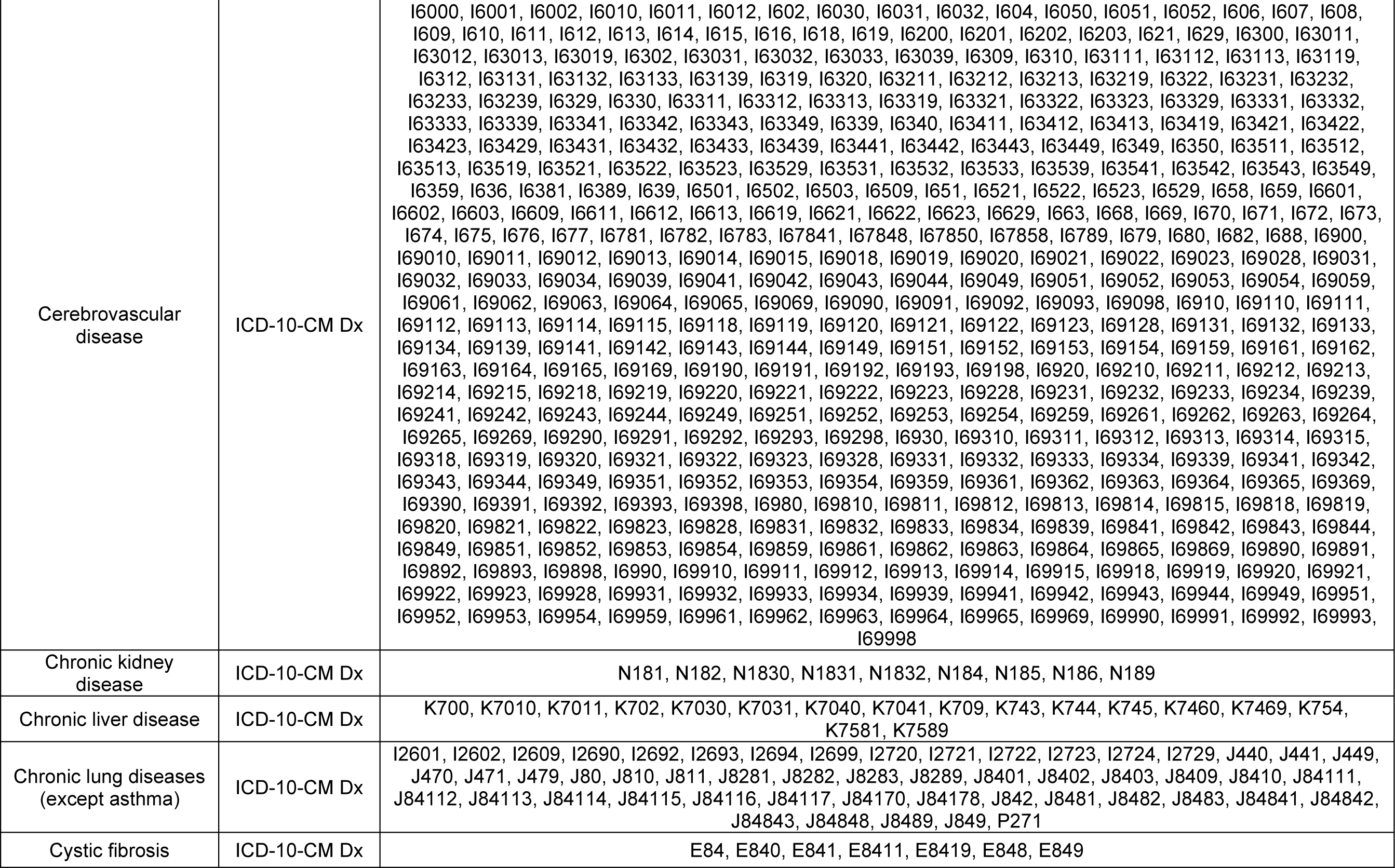
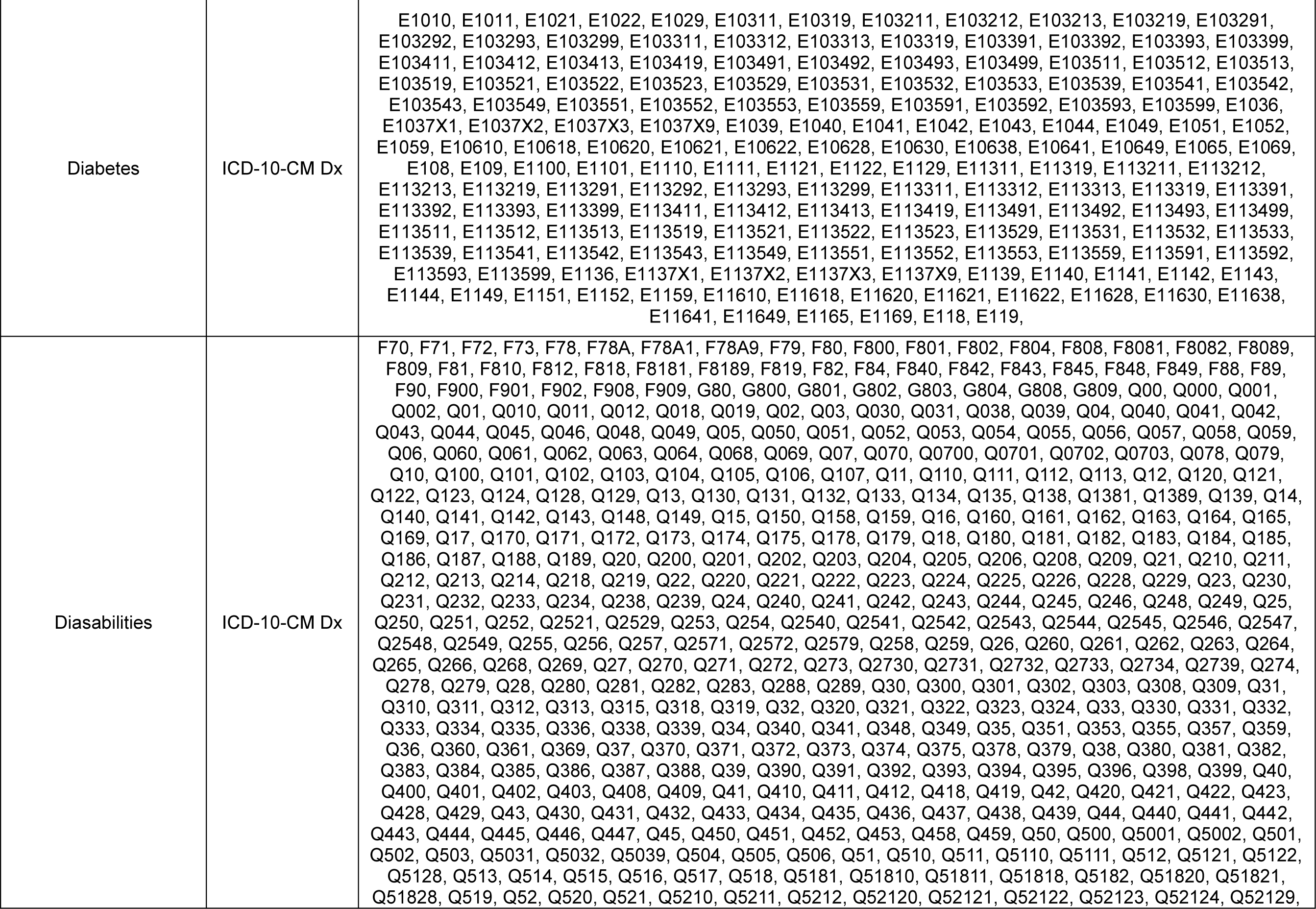
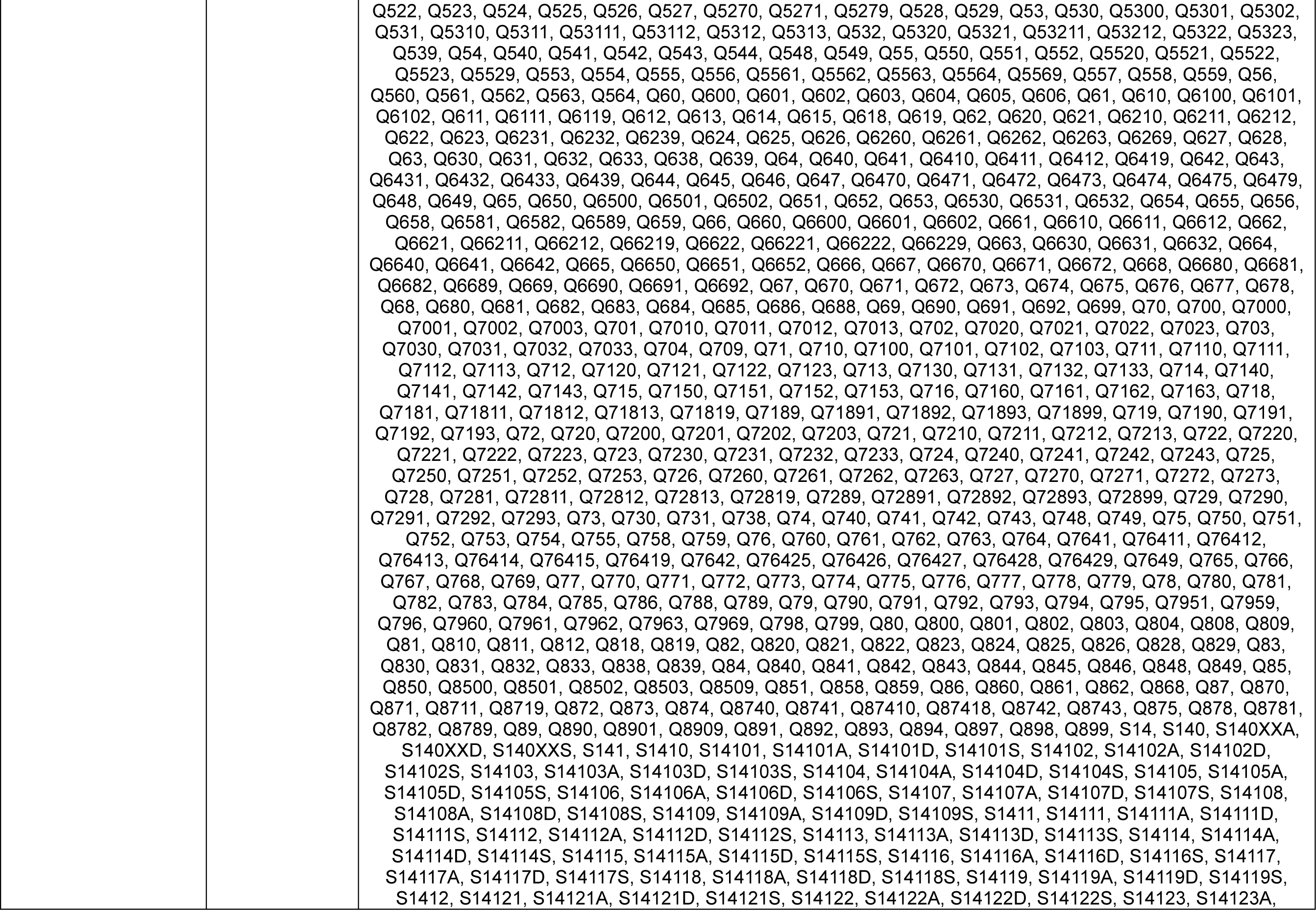
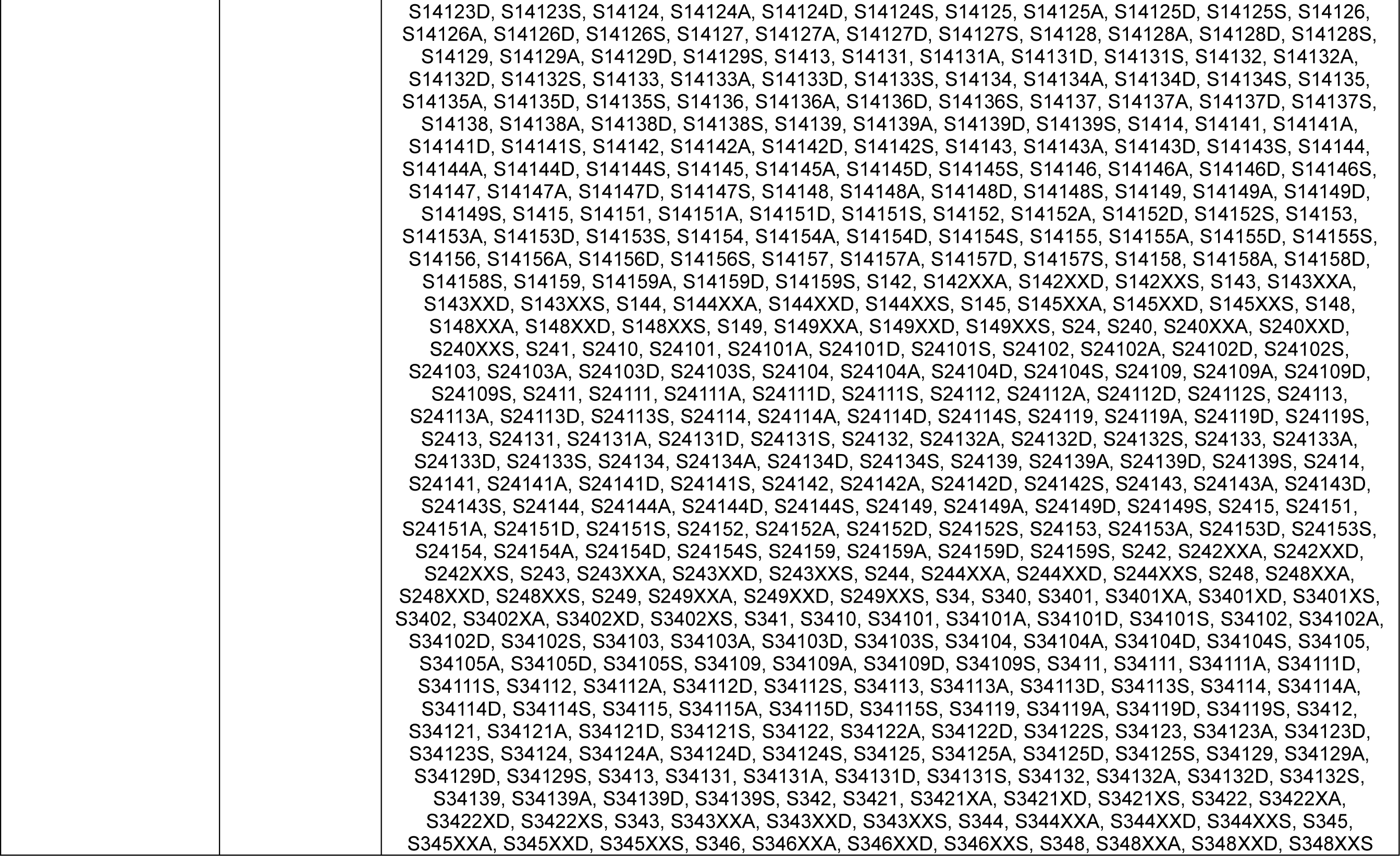
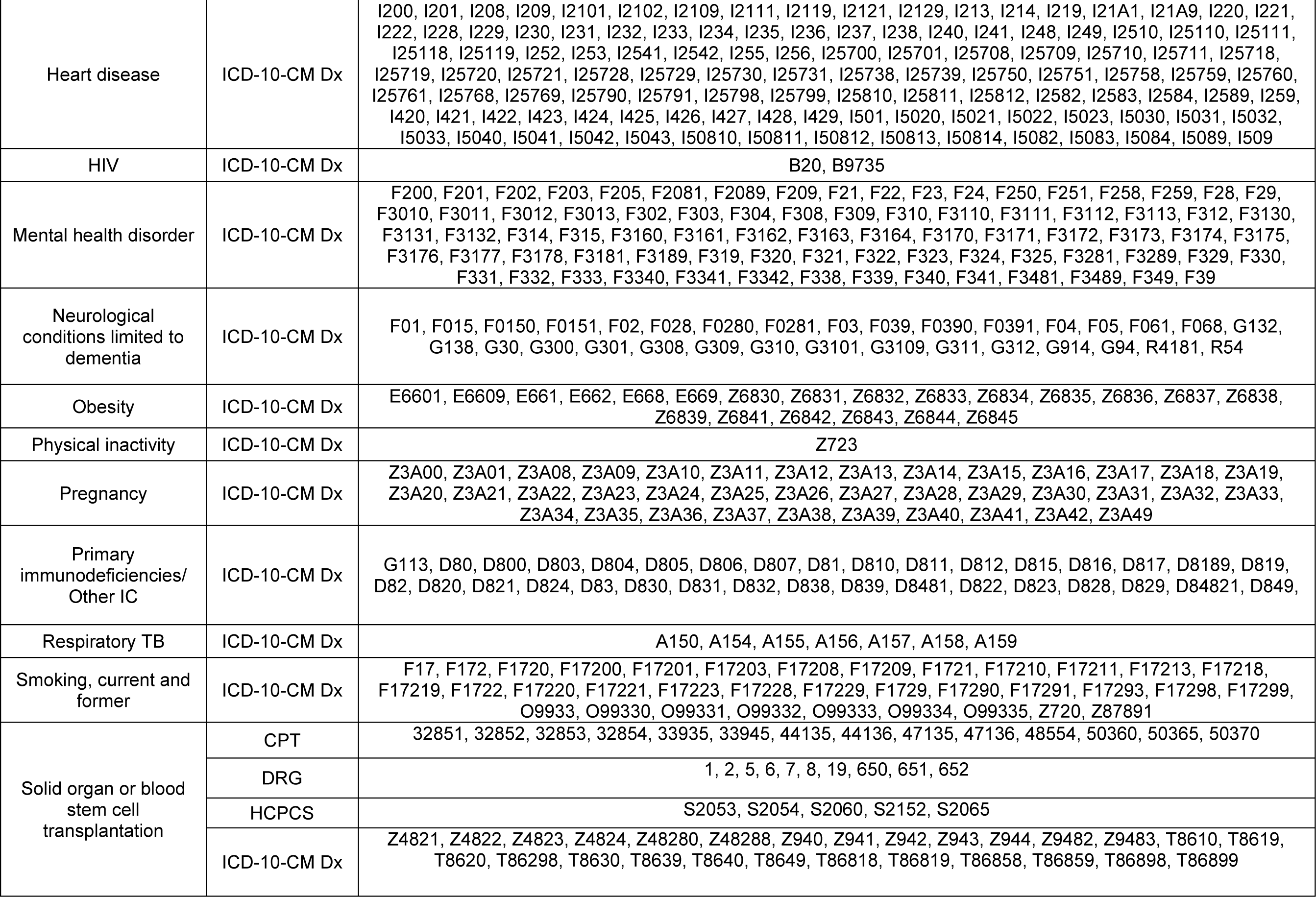
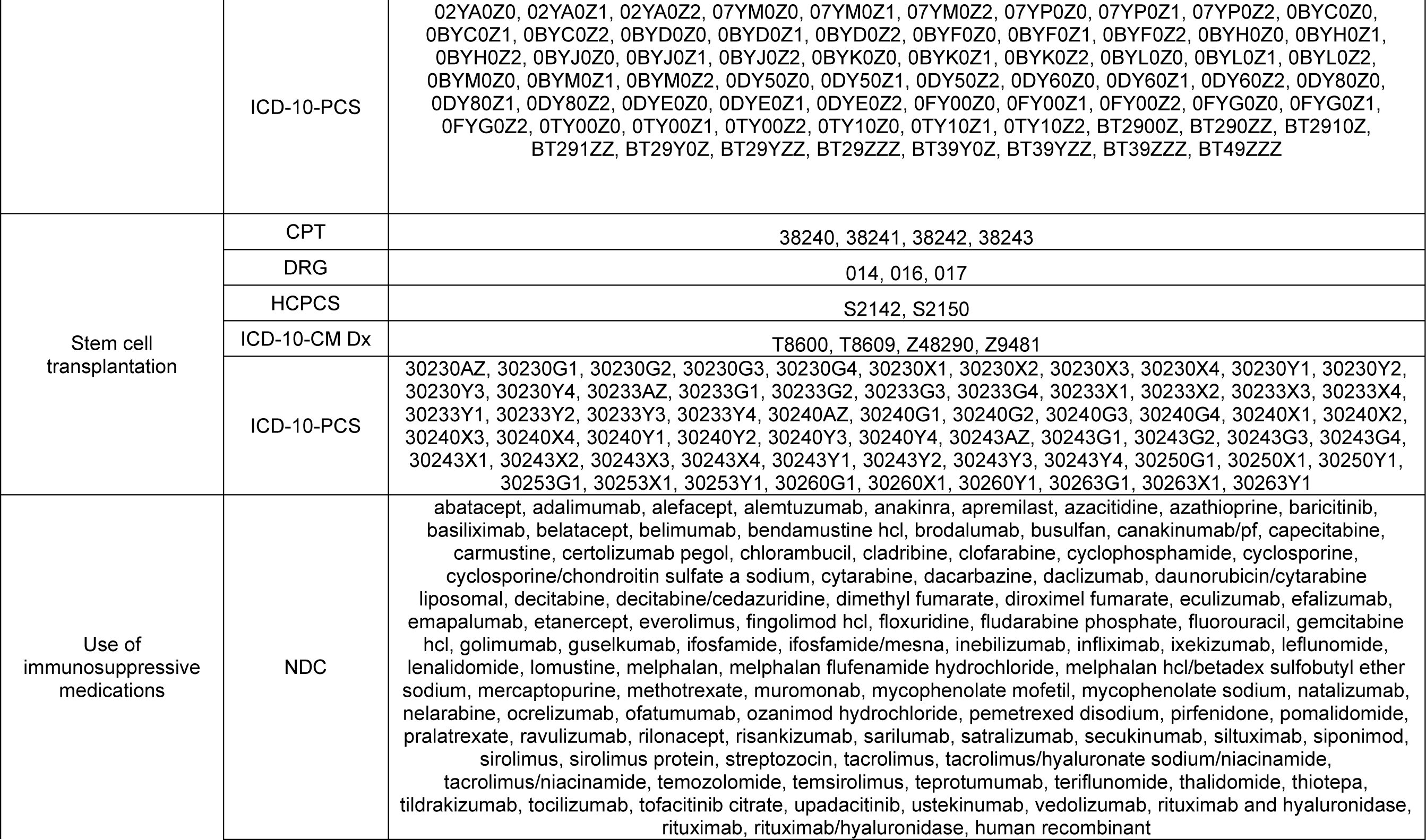
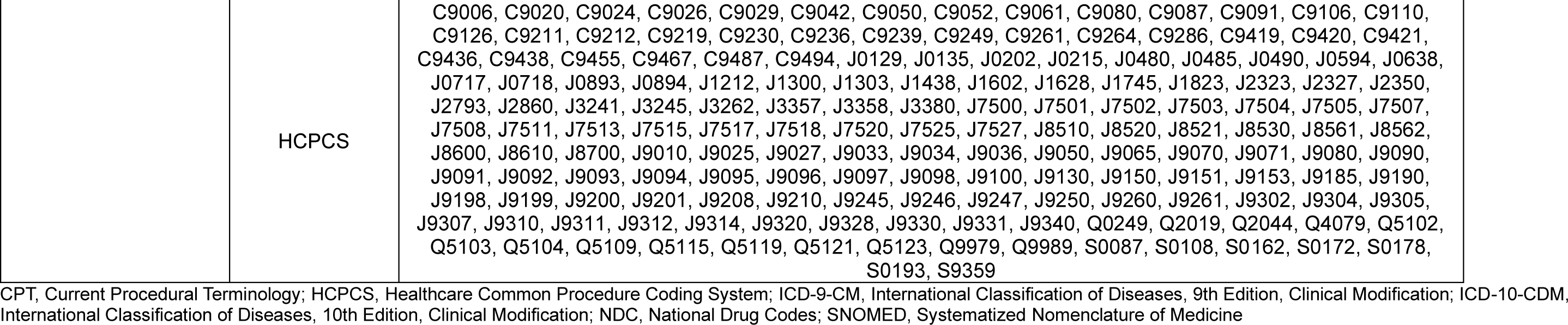
Code list for underlying medical conditions.

**Supplementary Table 2.**
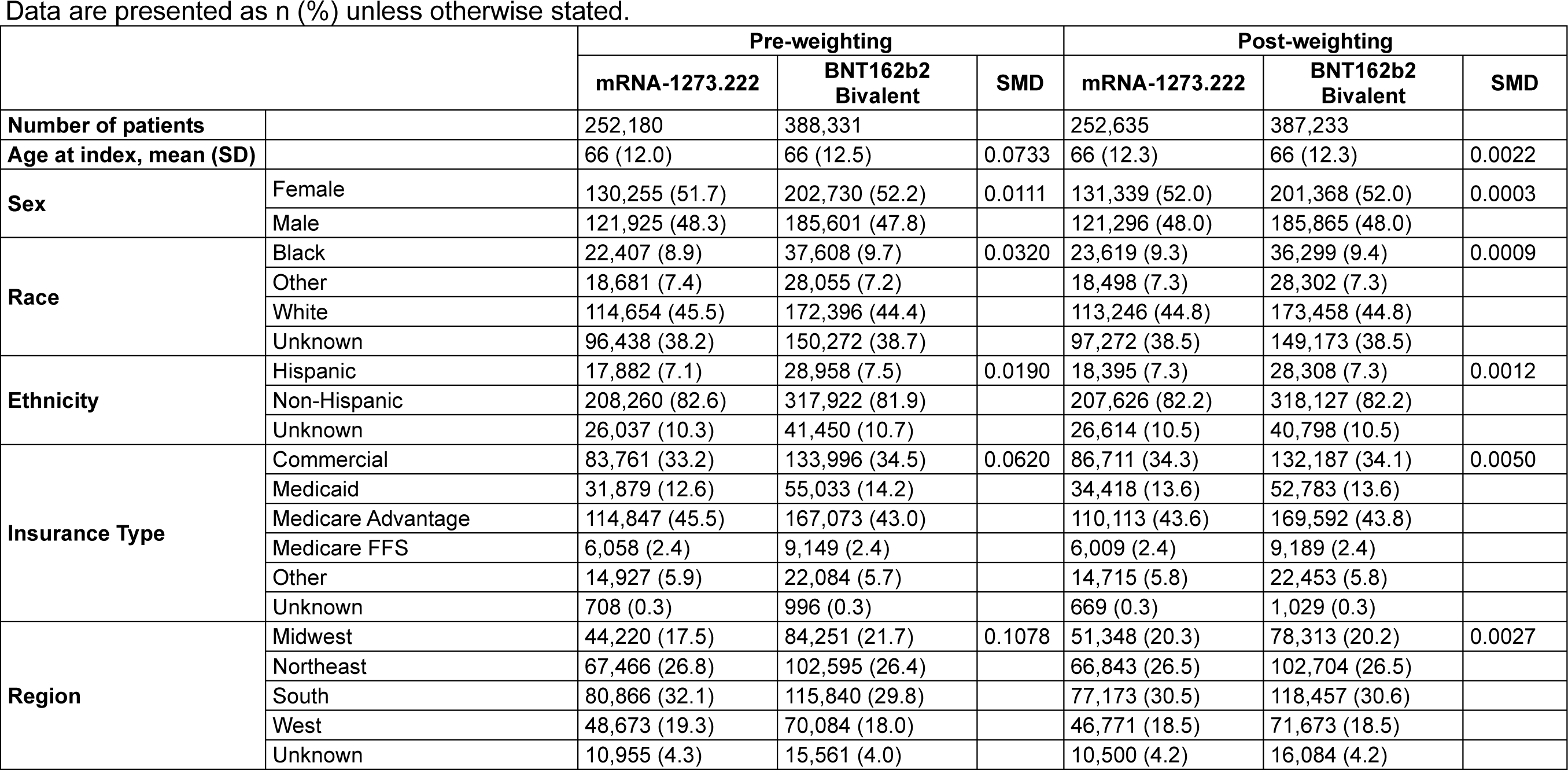
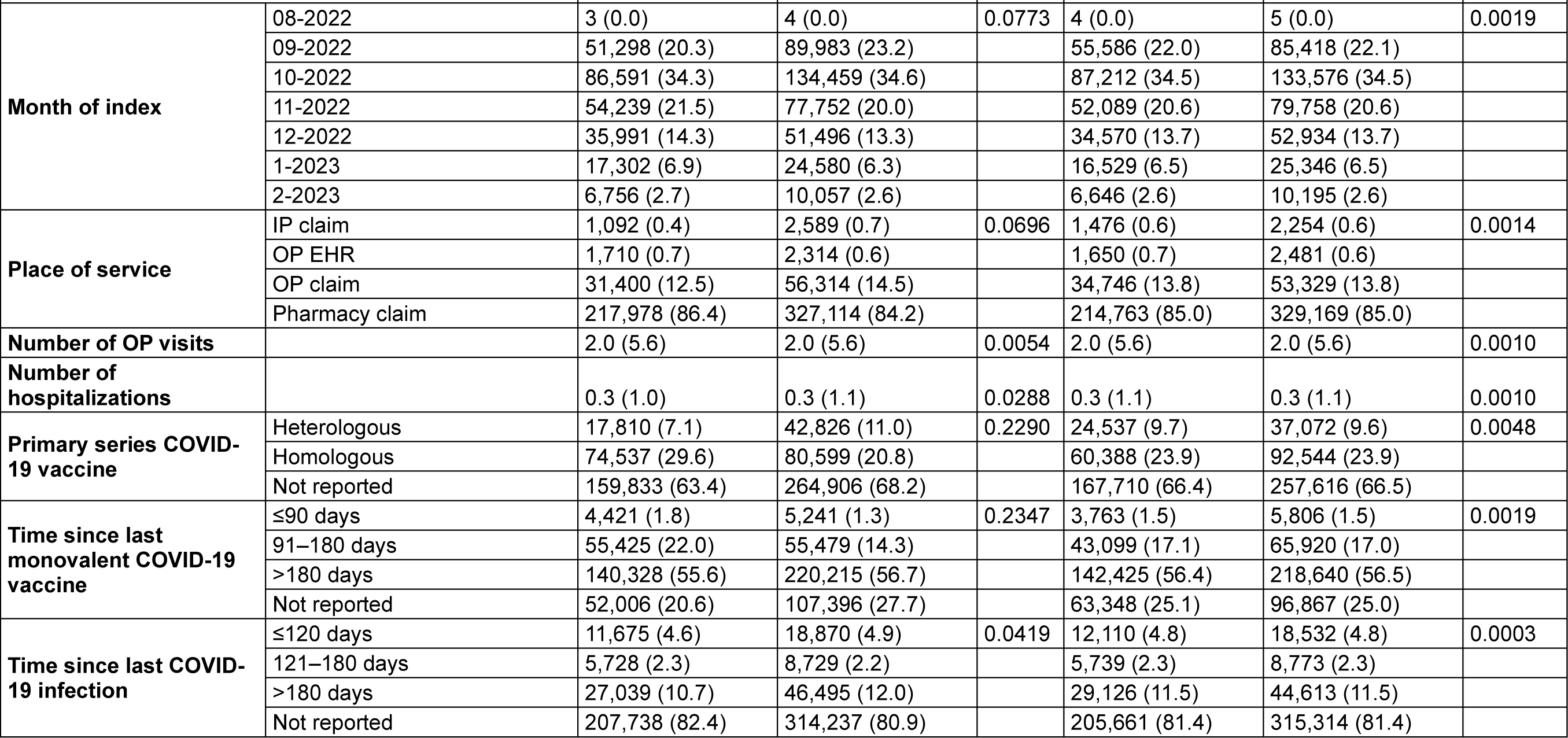
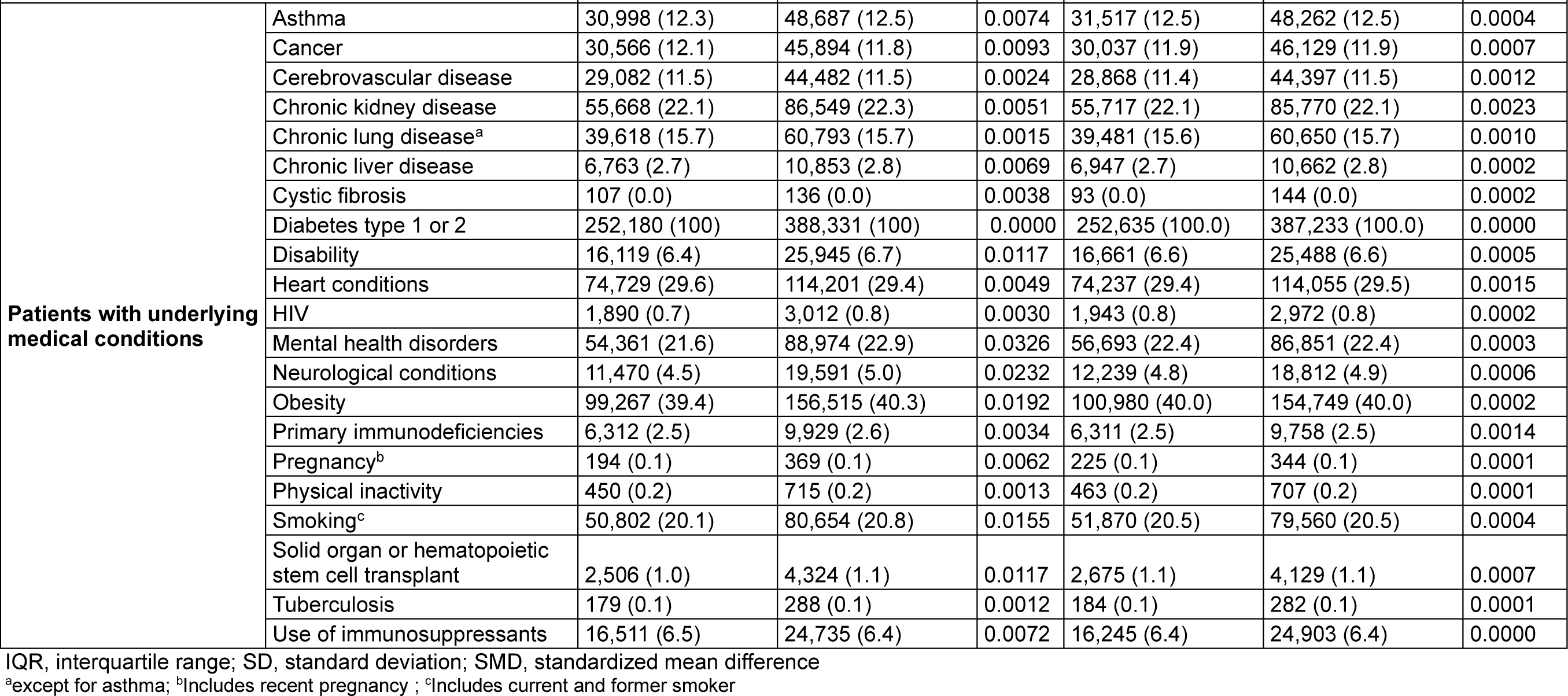
Baseline characteristics of the diabetic cohort.

Data are presented as n (%) unless otherwise stated.
|  |  | Pre-weighting |  |  | Post-weighting |  |  |
| --- | --- | --- | --- | --- | --- | --- | --- |
|  |  | mRNA-1273.222 | BNT162b2 Bivalent | SMD | mRNA-1273.222 | BNT162b2 Bivalent | SMD |
| **Number of patients** |  | 252,180 | 388,331 |  | 252,635 | 387,233 |  |
| **Age at index, mean (SD)** |  | 66 (12.0) | 66 (12.5) | 0.0733 | 66 (12.3) | 66 (12.3) | 0.0022 |
| **Sex** | Female | 130,255 (51.7) | 202,730 (52.2) | 0.0111 | 131,339 (52.0) | 201,368 (52.0) | 0.0003 |
|  | Male | 121,925 (48.3) | 185,601 (47.8) |  | 121,296 (48.0) | 185,865 (48.0) |  |
| **Race** | Black | 22,407 (8.9) | 37,608 (9.7) | 0.0320 | 23,619 (9.3) | 36,299 (9.4) | 0.0009 |
|  | Other | 18,681 (7.4) | 28,055 (7.2) |  | 18,498 (7.3) | 28,302 (7.3) |  |
|  | White | 114,654 (45.5) | 172,396 (44.4) |  | 113,246 (44.8) | 173,458 (44.8) |  |
|  | Unknown | 96,438 (38.2) | 150,272 (38.7) |  | 97,272 (38.5) | 149,173 (38.5) |  |
| **Ethnicity** | Hispanic | 17,882 (7.1) | 28,958 (7.5) | 0.0190 | 18,395 (7.3) | 28,308 (7.3) | 0.0012 |
|  | Non-Hispanic | 208,260 (82.6) | 317,922 (81.9) |  | 207,626 (82.2) | 318,127 (82.2) |  |
|  | Unknown | 26,037 (10.3) | 41,450 (10.7) |  | 26,614 (10.5) | 40,798 (10.5) |  |
| **Insurance Type** | Commercial | 83,761 (33.2) | 133,996 (34.5) | 0.0620 | 86,711 (34.3) | 132,187 (34.1) | 0.0050 |
|  | Medicaid | 31,879 (12.6) | 55,033 (14.2) |  | 34,418 (13.6) | 52,783 (13.6) |  |
|  | Medicare Advantage | 114,847 (45.5) | 167,073 (43.0) |  | 110,113 (43.6) | 169,592 (43.8) |  |
|  | Medicare FFS | 6,058 (2.4) | 9,149 (2.4) |  | 6,009 (2.4) | 9,189 (2.4) |  |
|  | Other | 14,927 (5.9) | 22,084 (5.7) |  | 14,715 (5.8) | 22,453 (5.8) |  |
|  | Unknown | 708 (0.3) | 996 (0.3) |  | 669 (0.3) | 1,029 (0.3) |  |
| **Region** | Midwest | 44,220 (17.5) | 84,251 (21.7) | 0.1078 | 51,348 (20.3) | 78,313 (20.2) | 0.0027 |
|  | Northeast | 67,466 (26.8) | 102,595 (26.4) |  | 66,843 (26.5) | 102,704 (26.5) |  |
|  | South | 80,866 (32.1) | 115,840 (29.8) |  | 77,173 (30.5) | 118,457 (30.6) |  |
|  | West | 48,673 (19.3) | 70,084 (18.0) |  | 46,771 (18.5) | 71,673 (18.5) |  |
|  | Unknown | 10,955 (4.3) | 15,561 (4.0) |  | 10,500 (4.2) | 16,084 (4.2) |  |
|  |  | mRNA-1273.222 | BNT162b2 Bivalent | SMD | mRNA-1273.222 | BNT162b2 Bivalent | SMD |
| Month of index | 08-2022 | 3 (0.0) | 4 (0.0) | 0.0773 | 4 (0.0) | 5 (0.0) | 0.0019 |
|  | 09-2022 | 51,298 (20.3) | 89,983 (23.2) |  | 55,586 (22.0) | 85,418 (22.1) |  |
|  | 10-2022 | 86,591 (34.3) | 134,459 (34.6) |  | 87,212 (34.5) | 133,576 (34.5) |  |
|  | 11-2022 | 54,239 (21.5) | 77,752 (20.0) |  | 52,089 (20.6) | 79,758 (20.6) |  |
|  | 12-2022 | 35,991 (14.3) | 51,496 (13.3) |  | 34,570 (13.7) | 52,934 (13.7) |  |
|  | 1-2023 | 17,302 (6.9) | 24,580 (6.3) |  | 16,529 (6.5) | 25,346 (6.5) |  |
|  | 2-2023 | 6,756 (2.7) | 10,057 (2.6) |  | 6,646 (2.6) | 10,195 (2.6) |  |
| Place of service | IP claim | 1,092 (0.4) | 2,589 (0.7) | 0.0696 | 1,476 (0.6) | 2,254 (0.6) | 0.0014 |
|  | OP EHR | 1,710 (0.7) | 2,314 (0.6) |  | 1,650 (0.7) | 2,481 (0.6) |  |
|  | OP claim | 31,400 (12.5) | 56,314 (14.5) |  | 34,746 (13.8) | 53,329 (13.8) |  |
|  | Pharmacy claim | 217,978 (86.4) | 327,114 (84.2) |  | 214,763 (85.0) | 329,169 (85.0) |  |
| Number of OP visits |  | 2.0 (5.6) | 2.0 (5.6) | 0.0054 | 2.0 (5.6) | 2.0 (5.6) | 0.0010 |
| Number of hospitalizations |  | 0.3 (1.0) | 0.3 (1.1) | 0.0288 | 0.3 (1.1) | 0.3 (1.1) | 0.0010 |
| Primary series COVID-19 vaccine | Heterologous | 17,810 (7.1) | 42,826 (11.0) | 0.2290 | 24,537 (9.7) | 37,072 (9.6) | 0.0048 |
|  | Homologous | 74,537 (29.6) | 80,599 (20.8) |  | 60,388 (23.9) | 92,544 (23.9) |  |
|  | Not reported | 159,833 (63.4) | 264,906 (68.2) |  | 167,710 (66.4) | 257,616 (66.5) |  |
| Time since last monovalent COVID-19 vaccine | ≤90 days | 4,421 (1.8) | 5,241 (1.3) | 0.2347 | 3,763 (1.5) | 5,806 (1.5) | 0.0019 |
|  | 91–180 days | 55,425 (22.0) | 55,479 (14.3) |  | 43,099 (17.1) | 65,920 (17.0) |  |
|  | >180 days | 140,328 (55.6) | 220,215 (56.7) |  | 142,425 (56.4) | 218,640 (56.5) |  |
|  | Not reported | 52,006 (20.6) | 107,396 (27.7) |  | 63,348 (25.1) | 96,867 (25.0) |  |
| Time since last COVID-19 infection | ≤120 days | 11,675 (4.6) | 18,870 (4.9) | 0.0419 | 12,110 (4.8) | 18,532 (4.8) | 0.0003 |
|  | 121–180 days | 5,728 (2.3) | 8,729 (2.2) |  | 5,739 (2.3) | 8,773 (2.3) |  |
|  | >180 days | 27,039 (10.7) | 46,495 (12.0) |  | 29,126 (11.5) | 44,613 (11.5) |  |
|  | Not reported | 207,738 (82.4) | 314,237 (80.9) |  | 205,661 (81.4) | 315,314 (81.4) |  |
|  |  | mRNA-1273.222 | BNT162b2<br>Bivalent | SMD | mRNA-1273.222 | BNT162b2<br>Bivalent | SMD |
| **Patients with underlying medical conditions** | Asthma | 30,998 (12.3) | 48,687 (12.5) | 0.0074 | 31,517 (12.5) | 48,262 (12.5) | 0.0004 |
|  | Cancer | 30,566 (12.1) | 45,894 (11.8) | 0.0093 | 30,037 (11.9) | 46,129 (11.9) | 0.0007 |
|  | Cerebrovascular disease | 29,082 (11.5) | 44,482 (11.5) | 0.0024 | 28,868 (11.4) | 44,397 (11.5) | 0.0012 |
|  | Chronic kidney disease | 55,668 (22.1) | 86,549 (22.3) | 0.0051 | 55,717 (22.1) | 85,770 (22.1) | 0.0023 |
|  | Chronic lung disease<sup>a</sup> | 39,618 (15.7) | 60,793 (15.7) | 0.0015 | 39,481 (15.6) | 60,650 (15.7) | 0.0010 |
|  | Chronic liver disease | 6,763 (2.7) | 10,853 (2.8) | 0.0069 | 6,947 (2.7) | 10,662 (2.8) | 0.0002 |
|  | Cystic fibrosis | 107 (0.0) | 136 (0.0) | 0.0038 | 93 (0.0) | 144 (0.0) | 0.0002 |
|  | Diabetes type 1 or 2 | 252,180 (100) | 388,331 (100) | 0.0000 | 252,635 (100.0) | 387,233 (100.0) | 0.0000 |
|  | Disability | 16,119 (6.4) | 25,945 (6.7) | 0.0117 | 16,661 (6.6) | 25,488 (6.6) | 0.0005 |
|  | Heart conditions | 74,729 (29.6) | 114,201 (29.4) | 0.0049 | 74,237 (29.4) | 114,055 (29.5) | 0.0015 |
|  | HIV | 1,890 (0.7) | 3,012 (0.8) | 0.0030 | 1,943 (0.8) | 2,972 (0.8) | 0.0002 |
|  | Mental health disorders | 54,361 (21.6) | 88,974 (22.9) | 0.0326 | 56,693 (22.4) | 86,851 (22.4) | 0.0003 |
|  | Neurological conditions | 11,470 (4.5) | 19,591 (5.0) | 0.0232 | 12,239 (4.8) | 18,812 (4.9) | 0.0006 |
|  | Obesity | 99,267 (39.4) | 156,515 (40.3) | 0.0192 | 100,980 (40.0) | 154,749 (40.0) | 0.0002 |
|  | Primary immunodeficiencies | 6,312 (2.5) | 9,929 (2.6) | 0.0034 | 6,311 (2.5) | 9,758 (2.5) | 0.0014 |
|  | Pregnancy<sup>b</sup> | 194 (0.1) | 369 (0.1) | 0.0062 | 225 (0.1) | 344 (0.1) | 0.0001 |
|  | Physical inactivity | 450 (0.2) | 715 (0.2) | 0.0013 | 463 (0.2) | 707 (0.2) | 0.0001 |
|  | Smoking<sup>c</sup> | 50,802 (20.1) | 80,654 (20.8) | 0.0155 | 51,870 (20.5) | 79,560 (20.5) | 0.0004 |
|  | Solid organ or hematopoietic stem cell transplant | 2,506 (1.0) | 4,324 (1.1) | 0.0117 | 2,675 (1.1) | 4,129 (1.1) | 0.0007 |
|  | Tuberculosis | 179 (0.1) | 288 (0.1) | 0.0012 | 184 (0.1) | 282 (0.1) | 0.0001 |
|  | Use of immunosuppressants | 16,511 (6.5) | 24,735 (6.4) | 0.0072 | 16,245 (6.4) | 24,903 (6.4) | 0.0000 |

IQR, interquartile range; SD, standard deviation; SMD, standardized mean difference
<sup>a</sup>except for asthma; <sup>b</sup>Includes recent pregnancy ; <sup>c</sup>Includes current and former smoker

**Supplementary Table 3.**
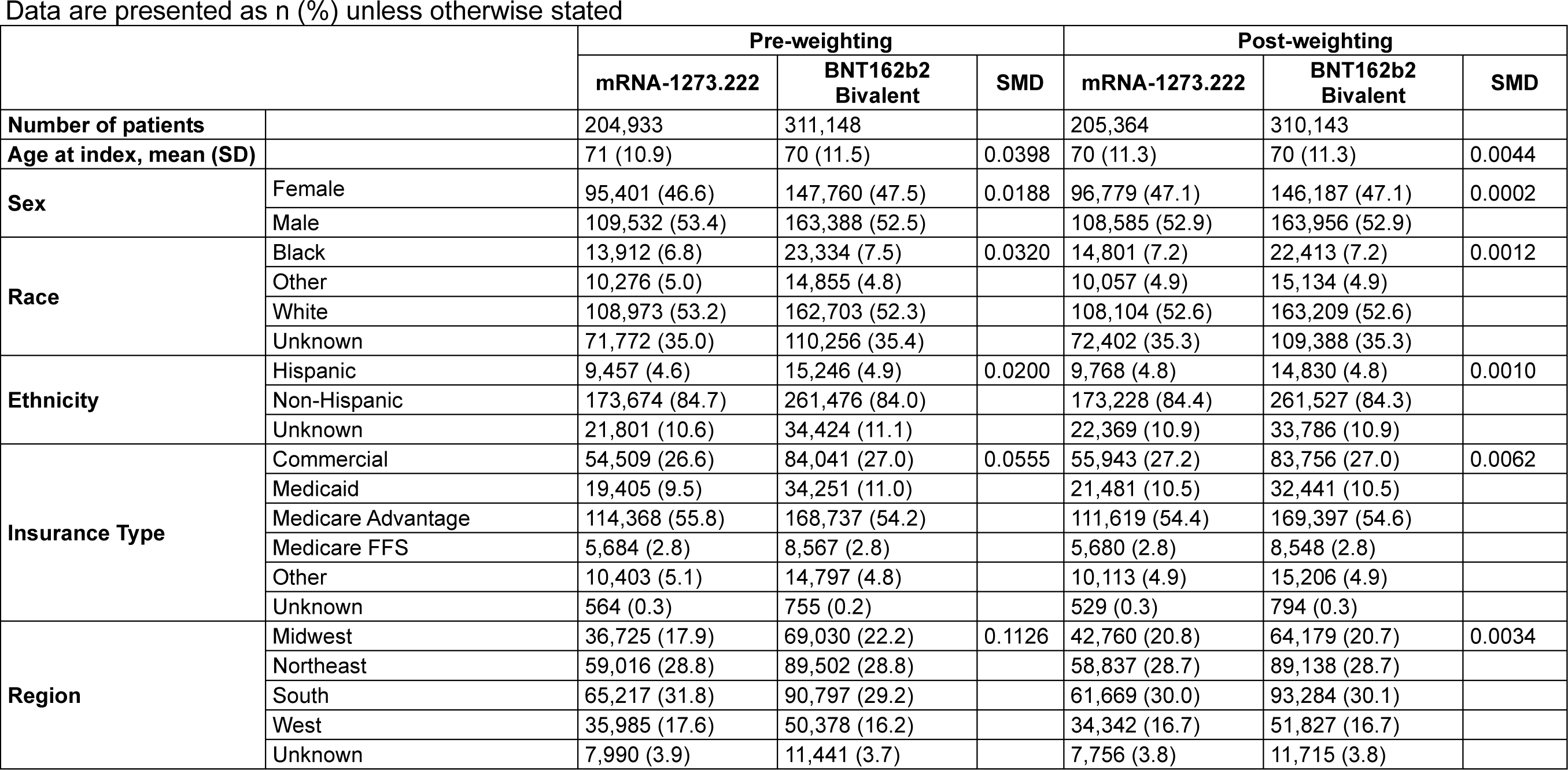
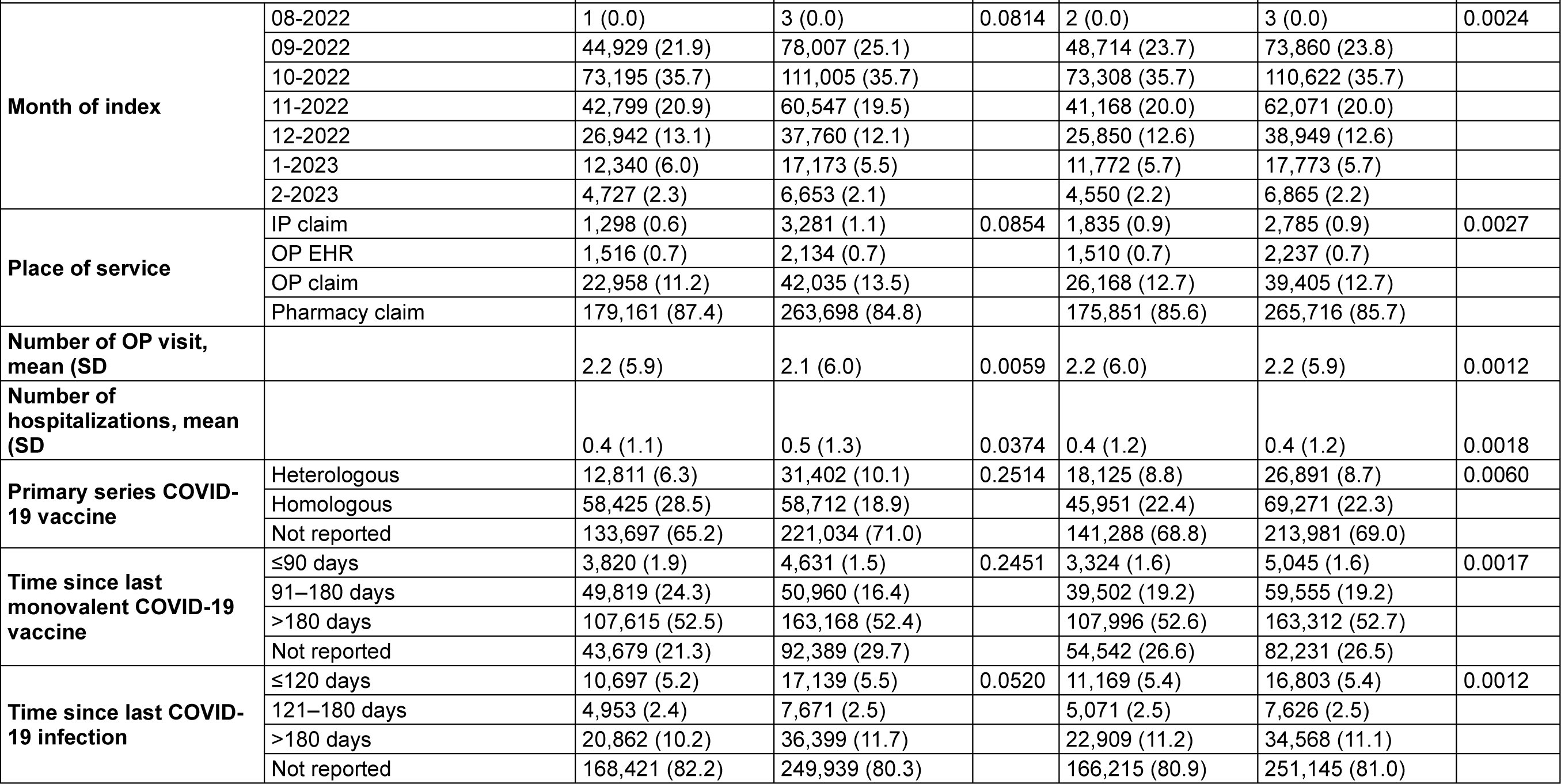
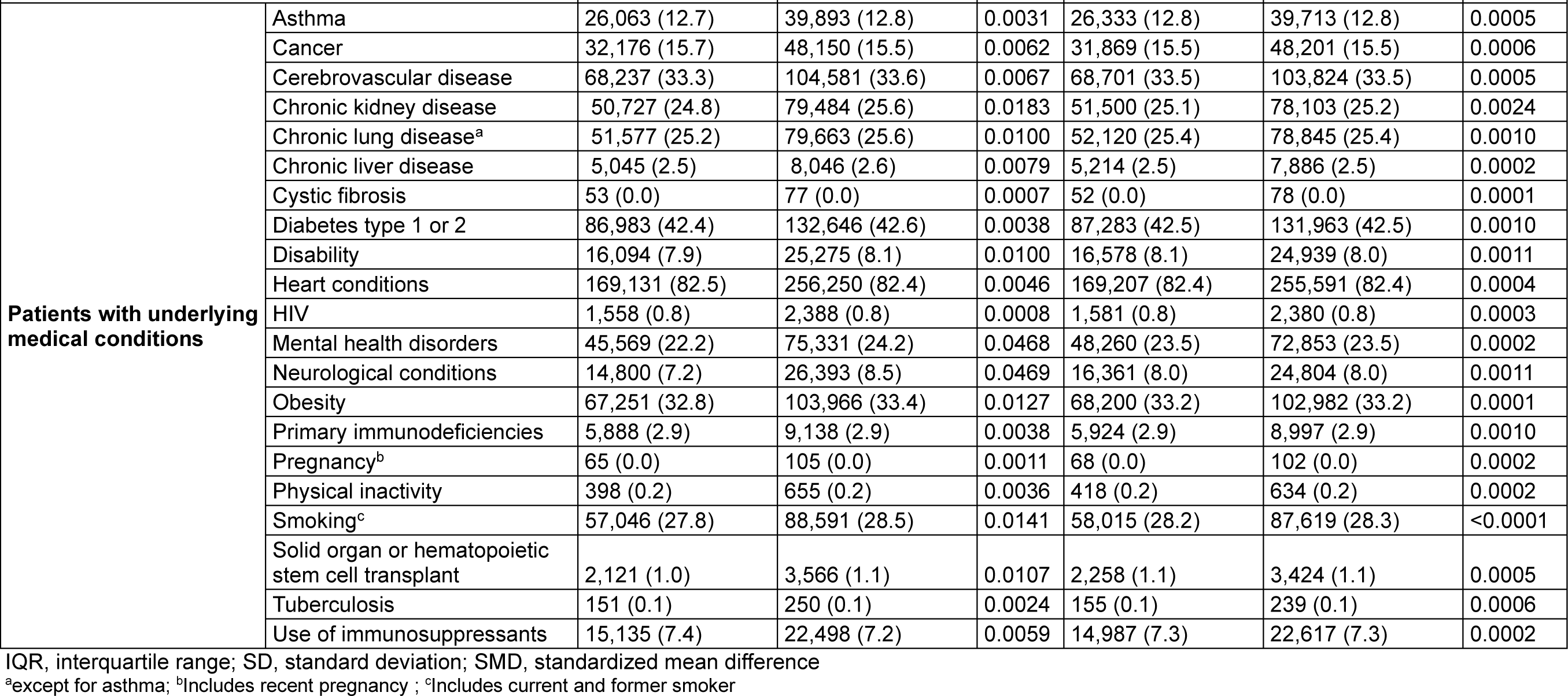
Baseline characteristics the cardiovascular cohort.

Data are presented as n (%) unless otherwise stated
|  |  | Pre-weighting |  |  | Post-weighting |  |  |
| --- | --- | --- | --- | --- | --- | --- | --- |
|  |  | mRNA-1273.222 | BNT162b2 Bivalent | SMD | mRNA-1273.222 | BNT162b2 Bivalent | SMD |
| **Number of patients** |  | 204,933 | 311,148 |  | 205,364 | 310,143 |  |
| **Age at index, mean (SD)** |  | 71 (10.9) | 70 (11.5) | 0.0398 | 70 (11.3) | 70 (11.3) | 0.0044 |
| **Sex** | Female | 95,401 (46.6) | 147,760 (47.5) | 0.0188 | 96,779 (47.1) | 146,187 (47.1) | 0.0002 |
|  | Male | 109,532 (53.4) | 163,388 (52.5) |  | 108,585 (52.9) | 163,956 (52.9) |  |
| **Race** | Black | 13,912 (6.8) | 23,334 (7.5) | 0.0320 | 14,801 (7.2) | 22,413 (7.2) | 0.0012 |
|  | Other | 10,276 (5.0) | 14,855 (4.8) |  | 10,057 (4.9) | 15,134 (4.9) |  |
|  | White | 108,973 (53.2) | 162,703 (52.3) |  | 108,104 (52.6) | 163,209 (52.6) |  |
|  | Unknown | 71,772 (35.0) | 110,256 (35.4) |  | 72,402 (35.3) | 109,388 (35.3) |  |
| **Ethnicity** | Hispanic | 9,457 (4.6) | 15,246 (4.9) | 0.0200 | 9,768 (4.8) | 14,830 (4.8) | 0.0010 |
|  | Non-Hispanic | 173,674 (84.7) | 261,476 (84.0) |  | 173,228 (84.4) | 261,527 (84.3) |  |
|  | Unknown | 21,801 (10.6) | 34,424 (11.1) |  | 22,369 (10.9) | 33,786 (10.9) |  |
| **Insurance Type** | Commercial | 54,509 (26.6) | 84,041 (27.0) | 0.0555 | 55,943 (27.2) | 83,756 (27.0) | 0.0062 |
|  | Medicaid | 19,405 (9.5) | 34,251 (11.0) |  | 21,481 (10.5) | 32,441 (10.5) |  |
|  | Medicare Advantage | 114,368 (55.8) | 168,737 (54.2) |  | 111,619 (54.4) | 169,397 (54.6) |  |
|  | Medicare FFS | 5,684 (2.8) | 8,567 (2.8) |  | 5,680 (2.8) | 8,548 (2.8) |  |
|  | Other | 10,403 (5.1) | 14,797 (4.8) |  | 10,113 (4.9) | 15,206 (4.9) |  |
|  | Unknown | 564 (0.3) | 755 (0.2) |  | 529 (0.3) | 794 (0.3) |  |
| **Region** | Midwest | 36,725 (17.9) | 69,030 (22.2) | 0.1126 | 42,760 (20.8) | 64,179 (20.7) | 0.0034 |
|  | Northeast | 59,016 (28.8) | 89,502 (28.8) |  | 58,837 (28.7) | 89,138 (28.7) |  |
|  | South | 65,217 (31.8) | 90,797 (29.2) |  | 61,669 (30.0) | 93,284 (30.1) |  |
|  | West | 35,985 (17.6) | 50,378 (16.2) |  | 34,342 (16.7) | 51,827 (16.7) |  |
|  | Unknown | 7,990 (3.9) | 11,441 (3.7) |  | 7,756 (3.8) | 11,715 (3.8) |  |
|  |  | mRNA-1273.222 | BNT162b2 Bivalent | SMD | mRNA-1273.222 | BNT162b2 Bivalent | SMD |
| Month of index | 08-2022 | 1 (0.0) | 3 (0.0) | 0.0814 | 2 (0.0) | 3 (0.0) | 0.0024 |
|  | 09-2022 | 44,929 (21.9) | 78,007 (25.1) |  | 48,714 (23.7) | 73,860 (23.8) |  |
|  | 10-2022 | 73,195 (35.7) | 111,005 (35.7) |  | 73,308 (35.7) | 110,622 (35.7) |  |
|  | 11-2022 | 42,799 (20.9) | 60,547 (19.5) |  | 41,168 (20.0) | 62,071 (20.0) |  |
|  | 12-2022 | 26,942 (13.1) | 37,760 (12.1) |  | 25,850 (12.6) | 38,949 (12.6) |  |
|  | 1-2023 | 12,340 (6.0) | 17,173 (5.5) |  | 11,772 (5.7) | 17,773 (5.7) |  |
|  | 2-2023 | 4,727 (2.3) | 6,653 (2.1) |  | 4,550 (2.2) | 6,865 (2.2) |  |
| Place of service | IP claim | 1,298 (0.6) | 3,281 (1.1) | 0.0854 | 1,835 (0.9) | 2,785 (0.9) | 0.0027 |
|  | OP EHR | 1,516 (0.7) | 2,134 (0.7) |  | 1,510 (0.7) | 2,237 (0.7) |  |
|  | OP claim | 22,958 (11.2) | 42,035 (13.5) |  | 26,168 (12.7) | 39,405 (12.7) |  |
|  | Pharmacy claim | 179,161 (87.4) | 263,698 (84.8) |  | 175,851 (85.6) | 265,716 (85.7) |  |
| Number of OP visit, mean (SD) |  | 2.2 (5.9) | 2.1 (6.0) | 0.0059 | 2.2 (6.0) | 2.2 (5.9) | 0.0012 |
| Number of hospitalizations, mean (SD) |  | 0.4 (1.1) | 0.5 (1.3) | 0.0374 | 0.4 (1.2) | 0.4 (1.2) | 0.0018 |
| Primary series COVID-19 vaccine | Heterologous | 12,811 (6.3) | 31,402 (10.1) | 0.2514 | 18,125 (8.8) | 26,891 (8.7) | 0.0060 |
|  | Homologous | 58,425 (28.5) | 58,712 (18.9) |  | 45,951 (22.4) | 69,271 (22.3) |  |
|  | Not reported | 133,697 (65.2) | 221,034 (71.0) |  | 141,288 (68.8) | 213,981 (69.0) |  |
| Time since last monovalent COVID-19 vaccine | ≤90 days | 3,820 (1.9) | 4,631 (1.5) | 0.2451 | 3,324 (1.6) | 5,045 (1.6) | 0.0017 |
|  | 91–180 days | 49,819 (24.3) | 50,960 (16.4) |  | 39,502 (19.2) | 59,555 (19.2) |  |
|  | >180 days | 107,615 (52.5) | 163,168 (52.4) |  | 107,996 (52.6) | 163,312 (52.7) |  |
|  | Not reported | 43,679 (21.3) | 92,389 (29.7) |  | 54,542 (26.6) | 82,231 (26.5) |  |
| Time since last COVID-19 infection | ≤120 days | 10,697 (5.2) | 17,139 (5.5) | 0.0520 | 11,169 (5.4) | 16,803 (5.4) | 0.0012 |
|  | 121–180 days | 4,953 (2.4) | 7,671 (2.5) |  | 5,071 (2.5) | 7,626 (2.5) |  |
|  | >180 days | 20,862 (10.2) | 36,399 (11.7) |  | 22,909 (11.2) | 34,568 (11.1) |  |
|  | Not reported | 168,421 (82.2) | 249,939 (80.3) |  | 166,215 (80.9) | 251,145 (81.0) |  |
|  |  | mRNA-1273.222 | BNT162b2 Bivalent | SMD | mRNA-1273.222 | BNT162b2 Bivalent | SMD |
| **Patients with underlying medical conditions** | Asthma | 26,063 (12.7) | 39,893 (12.8) | 0.0031 | 26,333 (12.8) | 39,713 (12.8) | 0.0005 |
|  | Cancer | 32,176 (15.7) | 48,150 (15.5) | 0.0062 | 31,869 (15.5) | 48,201 (15.5) | 0.0006 |
|  | Cerebrovascular disease | 68,237 (33.3) | 104,581 (33.6) | 0.0067 | 68,701 (33.5) | 103,824 (33.5) | 0.0005 |
|  | Chronic kidney disease | 50,727 (24.8) | 79,484 (25.6) | 0.0183 | 51,500 (25.1) | 78,103 (25.2) | 0.0024 |
|  | Chronic lung disease<sup>a</sup> | 51,577 (25.2) | 79,663 (25.6) | 0.0100 | 52,120 (25.4) | 78,845 (25.4) | 0.0010 |
|  | Chronic liver disease | 5,045 (2.5) | 8,046 (2.6) | 0.0079 | 5,214 (2.5) | 7,886 (2.5) | 0.0002 |
|  | Cystic fibrosis | 53 (0.0) | 77 (0.0) | 0.0007 | 52 (0.0) | 78 (0.0) | 0.0001 |
|  | Diabetes type 1 or 2 | 86,983 (42.4) | 132,646 (42.6) | 0.0038 | 87,283 (42.5) | 131,963 (42.5) | 0.0010 |
|  | Disability | 16,094 (7.9) | 25,275 (8.1) | 0.0100 | 16,578 (8.1) | 24,939 (8.0) | 0.0011 |
|  | Heart conditions | 169,131 (82.5) | 256,250 (82.4) | 0.0046 | 169,207 (82.4) | 255,591 (82.4) | 0.0004 |
|  | HIV | 1,558 (0.8) | 2,388 (0.8) | 0.0008 | 1,581 (0.8) | 2,380 (0.8) | 0.0003 |
|  | Mental health disorders | 45,569 (22.2) | 75,331 (24.2) | 0.0468 | 48,260 (23.5) | 72,853 (23.5) | 0.0002 |
|  | Neurological conditions | 14,800 (7.2) | 26,393 (8.5) | 0.0469 | 16,361 (8.0) | 24,804 (8.0) | 0.0011 |
|  | Obesity | 67,251 (32.8) | 103,966 (33.4) | 0.0127 | 68,200 (33.2) | 102,982 (33.2) | 0.0001 |
|  | Primary immunodeficiencies | 5,888 (2.9) | 9,138 (2.9) | 0.0038 | 5,924 (2.9) | 8,997 (2.9) | 0.0010 |
|  | Pregnancy<sup>b</sup> | 65 (0.0) | 105 (0.0) | 0.0011 | 68 (0.0) | 102 (0.0) | 0.0002 |
|  | Physical inactivity | 398 (0.2) | 655 (0.2) | 0.0036 | 418 (0.2) | 634 (0.2) | 0.0002 |
|  | Smoking<sup>c</sup> | 57,046 (27.8) | 88,591 (28.5) | 0.0141 | 58,015 (28.2) | 87,619 (28.3) | <0.0001 |
|  | Solid organ or hematopoietic stem cell transplant | 2,121 (1.0) | 3,566 (1.1) | 0.0107 | 2,258 (1.1) | 3,424 (1.1) | 0.0005 |
|  | Tuberculosis | 151 (0.1) | 250 (0.1) | 0.0024 | 155 (0.1) | 239 (0.1) | 0.0006 |
|  | Use of immunosuppressants | 15,135 (7.4) | 22,498 (7.2) | 0.0059 | 14,987 (7.3) | 22,617 (7.3) | 0.0002 |

IQR, interquartile range; SD, standard deviation; SMD, standardized mean difference
<sup>a</sup>except for asthma; <sup>b</sup>Includes recent pregnancy ; <sup>c</sup>Includes current and former smoker

**Supplementary Table 4.**
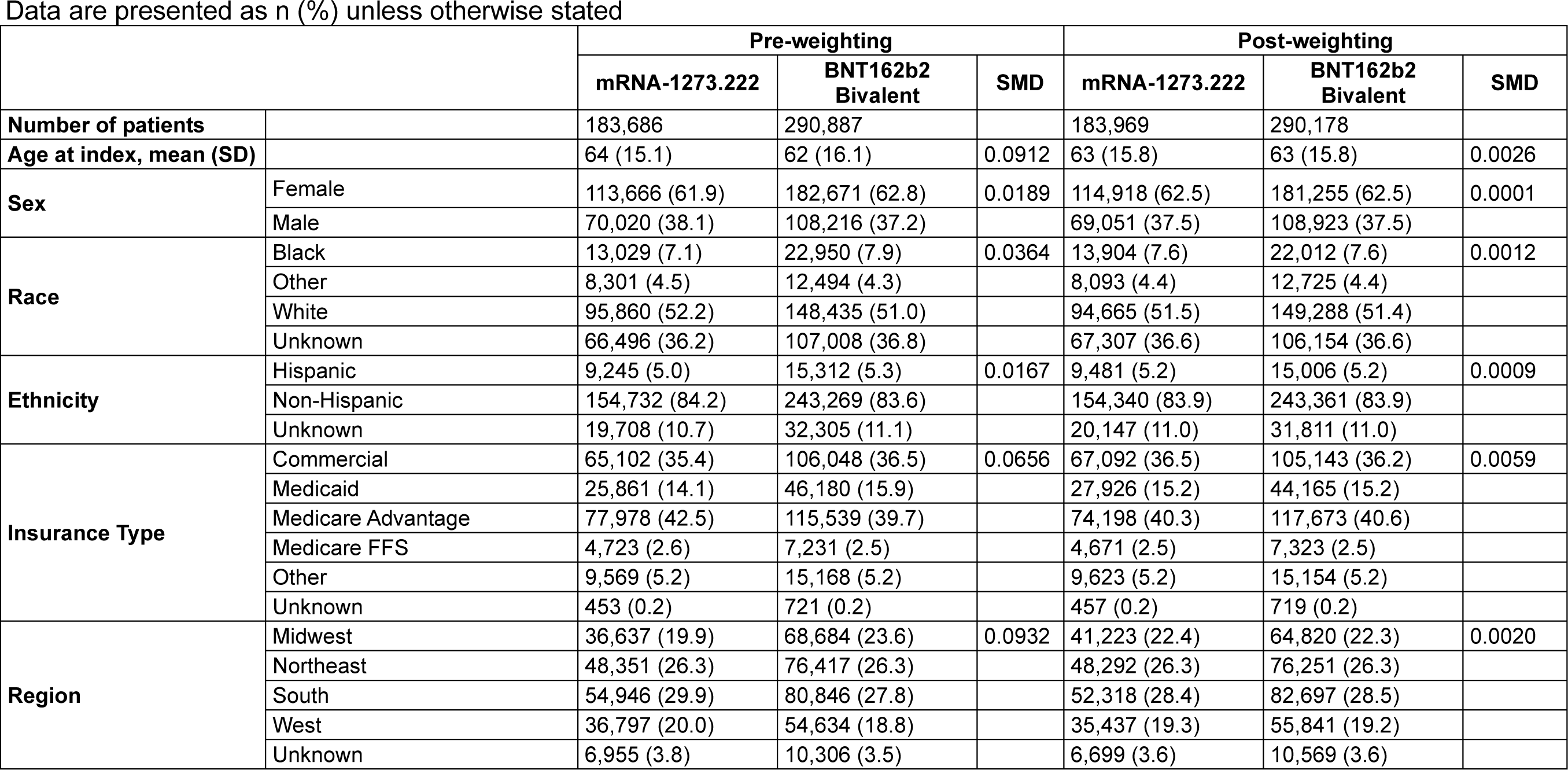
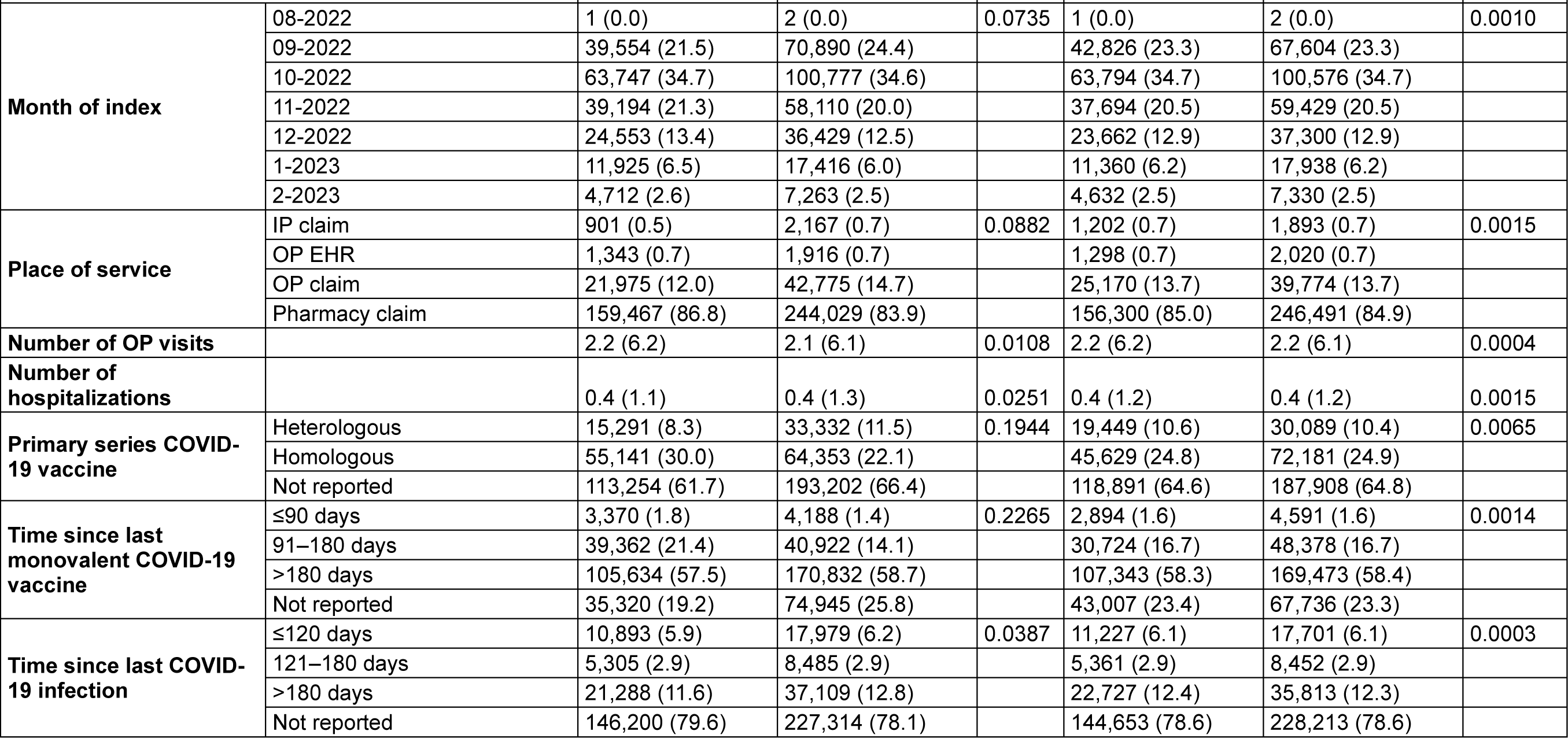
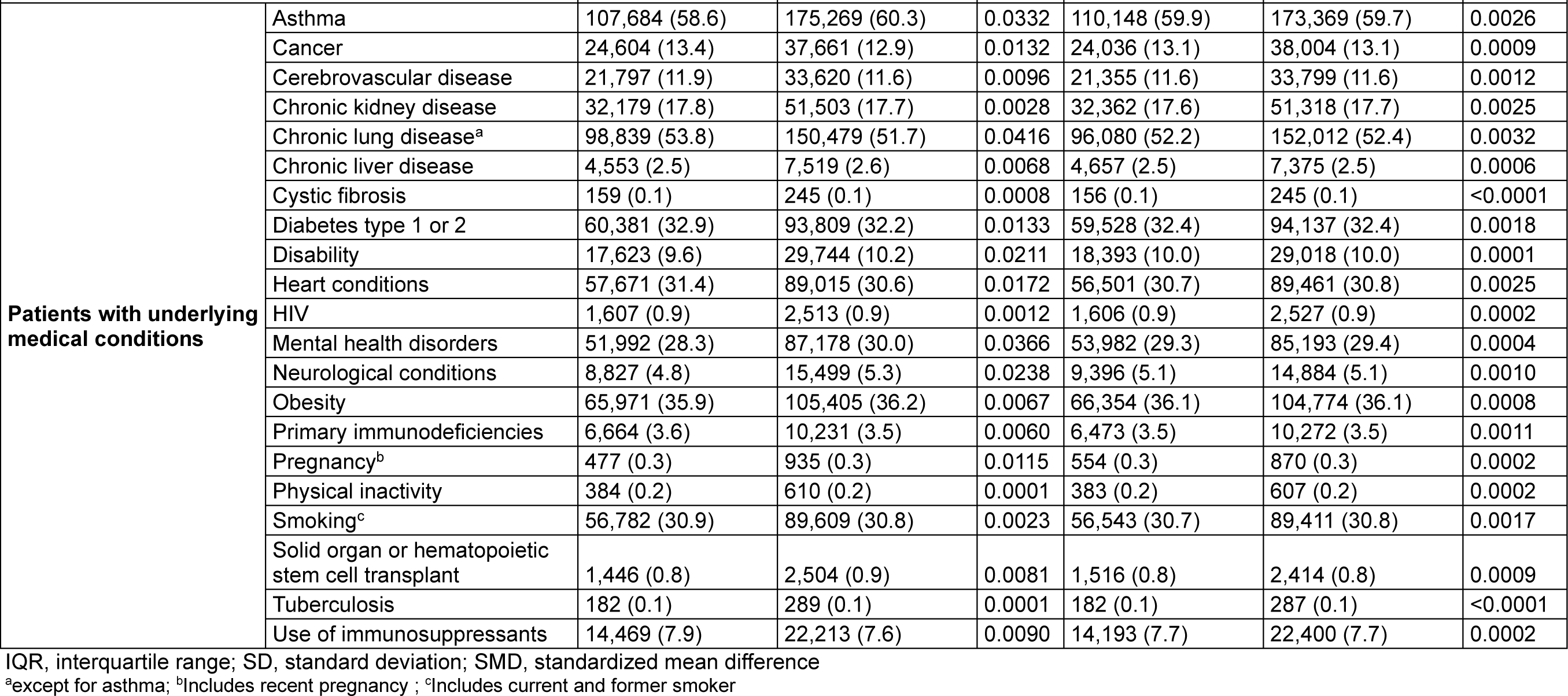
Baseline characteristics of the chronic lung disease cohort.

**Supplementary Table 5.** Baseline characteristics of the immunocompromised cohort.

Data are presented as n (%) unless otherwise stated
|  |  | Pre-weighting |  |  | Post-weighting |  |  |
| --- | --- | --- | --- | --- | --- | --- | --- |
|  |  | mRNA-1273.222 | BNT162b2 Bivalent | SMD | mRNA-1273.222 | BNT162b2 Bivalent | SMD |
| **Number of patients** |  | 169,185 | 258,467 |  | 169,509 | 257,751 |  |
| **Age at index, mean (SD)** |  | 66 (13.1) | 65 (13.6) | 0.0763 | 65 (13.4) | 65 (13.4) | 0.0028 |
| **Sex** | Female | 93,938 (55.5) | 145,949 (56.5) | 0.0190 | 95,086 (56.1) | 144,623 (56.1) | 0.0003 |
|  | Male | 75,247 (44.5) | 112,518 (43.5) |  | 74,423 (43.9) | 113,128 (43.9) |  |
| **Race** | Black | 10,464 (6.2) | 18,188 (7.0) | 0.0408 | 11,311 (6.7) | 17,284 (6.7) | 0.0017 |
|  | Other | 8,572 (5.1) | 12,286 (4.8) |  | 8,291 (4.9) | 12,585 (4.9) |  |
|  | White | 89,479 (52.9) | 133,780 (51.8) |  | 88,531 (52.2) | 134,517 (52.2) |  |
|  | Unknown | 60,670 (35.9) | 94,213 (36.5) |  | 61,376 (36.2) | 93,365 (36.2) |  |
| **Ethnicity** | Hispanic | 7,823 (4.6) | 12,027 (4.7) | 0.0159 | 7,824 (4.6) | 11,939 (4.6) | 0.0006 |
|  | Non-Hispanic | 142,195 (84.0) | 215,861 (83.5) |  | 141,982 (83.8) | 215,836 (83.7) |  |
|  | Unknown | 19,167 (11.3) | 30,576 (11.8) |  | 19,704 (11.6) | 29,976 (11.6) |  |
| **Insurance Type** | Commercial | 64,096 (37.9) | 101,719 (39.4) | 0.0576 | 66,475 (39.2) | 100,451 (39.0) | 0.0058 |
|  | Medicaid | 13,640 (8.1) | 23,449 (9.1) |  | 14,719 (8.7) | 22,431 (8.7) |  |
|  | Medicare Advantage | 76,348 (45.1) | 110,638 (42.8) |  | 73,265 (43.2) | 112,072 (43.5) |  |
|  | Medicare FFS | 3,410 (2.0) | 4,570 (1.8) |  | 3,156 (1.9) | 4,782 (1.9) |  |
|  | Other | 11,262 (6.7) | 17,468 (6.8) |  | 11,477 (6.8) | 17,382 (6.7) |  |
|  | Unknown | 429 (0.3) | 623 (0.2) |  | 417 (0.2) | 633 (0.2) |  |
| **Region** | Midwest | 30,527 (18.0) | 56,105 (21.7) | 0.0951 | 34,769 (20.5) | 52,634 (20.4) | 0.0025 |
|  | Northeast | 45,788 (27.1) | 69,310 (26.8) |  | 45,502 (26.8) | 69,309 (26.9) |  |
|  | South | 51,420 (30.4) | 73,884 (28.6) |  | 49,379 (29.1) | 75,237 (29.2) |  |
|  | West | 35,446 (21.0) | 50,447 (19.5) |  | 34,010 (20.1) | 51,681 (20.1) |  |
|  | Unknown | 6,004 (3.5) | 8,721 (3.4) |  | 5,849 (3.5) | 8,890 (3.4) |  |
|  |  | mRNA-1273.222 | BNT162b2 Bivalent | SMD | mRNA-1273.222 | BNT162b2 Bivalent | SMD |
| Month of index | 08-2022 | 1 (0.0) | 3 (0.0) | 0.0689 | 2 (0.0) | 3 (0.0) | 0.0010 |
|  | 09-2022 | 39,363 (23.3) | 66,977 (25.9) |  | 42,141 (24.9) | 64,151 (24.9) |  |
|  | 10-2022 | 59,500 (35.2) | 91,220 (35.3) |  | 59,780 (35.3) | 90,820 (35.2) |  |
|  | 11-2022 | 35,176 (20.8) | 50,397 (19.5) |  | 33,942 (20.0) | 51,557 (20.0) |  |
|  | 12-2022 | 21,822 (12.9) | 30,902 (12.0) |  | 20,875 (12.3) | 31,757 (12.3) |  |
|  | 1-2023 | 9,618 (5.7) | 13,644 (5.3) |  | 9,198 (5.4) | 14,014 (5.4) |  |
|  | 2-2023 | 3,705 (2.2) | 5,324 (2.1) |  | 3,572 (2.1) | 5,449 (2.1) |  |
| Place of service | IP claim | 413 (0.2) | 951 (0.4) | 0.0683 | 539 (0.3) | 829 (0.3) | 0.0012 |
|  | OP EHR | 1,090 (0.6) | 1,610 (0.6) |  | 1,095 (0.6) | 1,649 (0.6) |  |
|  | OP claim | 17,354 (10.3) | 31,702 (12.3) |  | 19,561 (11.5) | 29,749 (11.5) |  |
|  | Pharmacy claim | 150,328 (88.9) | 224,204 (86.7) |  | 148,313 (87.5) | 225,524 (87.5) |  |
| Number of OP visits |  | 2.1 (5.7) | 2.0 (5.6) | 0.0115 | 2.0 (5.7) | 2.0 (5.6) | 0.0008 |
| Number of hospitalizations |  | 0.3 (0.9) | 0.3 (1.0) | 0.0234 | 0.3 (1.0) | 0.3 (1.0) | 0.0012 |
| Primary series COVID-19 vaccine | Heterologous | 12,436 (7.4) | 26,239 (10.2) | 0.2028 | 15,885 (9.4) | 23,649 (9.2) | 0.0066 |
|  | Homologous | 49,937 (29.5) | 54,850 (21.2) |  | 40,900 (24.1) | 62,249 (24.2) |  |
|  | Not reported | 106,812 (63.1) | 177,378 (68.6) |  | 112,724 (66.5) | 171,853 (66.7) |  |
| Time since last monovalent COVID-19 vaccine | ≤90 days | 3,079 (1.8) | 3,676 (1.4) | 0.2498 | 2,653 (1.6) | 4,054 (1.6) | 0.0011 |
|  | 91–180 days | 41,132 (24.3) | 41,064 (15.9) |  | 32,195 (19.0) | 48,836 (18.9) |  |
|  | >180 days | 93,746 (55.4) | 146,846 (56.8) |  | 95,584 (56.4) | 145,447 (56.4) |  |
|  | Not reported | 31,228 (18.5) | 66,881 (25.9) |  | 39,077 (23.1) | 59,414 (23.1) |  |
| Time since last COVID-19 infection | ≤120 days | 7,844 (4.6) | 12,398 (4.8) | 0.0300 | 8,062 (4.8) | 12,248 (4.8) | 0.0005 |
|  | 121–180 days | 4,075 (2.4) | 6,302 (2.4) |  | 4,127 (2.4) | 6,270 (2.4) |  |
|  | >180 days | 15,539 (9.2) | 25,877 (10.0) |  | 16,483 (9.7) | 25,045 (9.7) |  |
|  | Not reported | 141,727 (83.8) | 213,890 (82.8) |  | 140,838 (83.1) | 214,188 (83.1) |  |
|  |  | mRNA-1273.222 | BNT162b2 Bivalent | SMD | mRNA-1273.222 | BNT162b2 Bivalent | SMD |
| **Patients with underlying medical conditions** | Asthma | 20,039 (11.8) | 31,209 (12.1) | 0.0071 | 20,347 (12.0) | 30,931 (12.0) | 0.0001 |
|  | Cancer | 99,704 (58.9) | 151,325 (58.5) | 0.0078 | 99,361 (58.6) | 151,212 (58.7) | 0.0010 |
|  | Cerebrovascular disease | 16,402 (9.7) | 24,583 (9.5) | 0.0062 | 16,127 (9.5) | 24,622 (9.6) | 0.0013 |
|  | Chronic kidney disease | 29,405 (17.4) | 45,001 (17.4) | 0.0008 | 29,223 (17.2) | 44,691 (17.3) | 0.0026 |
|  | Chronic lung disease<sup>a</sup> | 27,936 (16.5) | 41,819 (16.2) | 0.0090 | 27,489 (16.2) | 41,951 (16.3) | 0.0016 |
|  | Chronic liver disease | 4,246 (2.5) | 6,803 (2.6) | 0.0077 | 4,350 (2.6) | 6,652 (2.6) | 0.0009 |
|  | Cystic fibrosis | 95 (0.1) | 131 (0.1) | 0.0024 | 88 (0.1) | 135 (0.1) | 0.0001 |
|  | Diabetes type 1 or 2 | 49,366 (29.2) | 74,223 (28.7) | 0.0102 | 48,735 (28.8) | 74,326 (28.8) | 0.0019 |
|  | Disability | 12,658 (7.5) | 19,945 (7.7) | 0.0089 | 12,944 (7.6) | 19,668 (7.6) | 0.0002 |
|  | Heart conditions | 40,636 (24.0) | 60,807 (23.5) | 0.0116 | 39,987 (23.6) | 60,993 (23.7) | 0.0017 |
|  | HIV | 7,327 (4.3) | 11,394 (4.4) | 0.0038 | 7,467 (4.4) | 11,324 (4.4) | 0.0006 |
|  | Mental health disorders | 33,435 (19.8) | 54,152 (21.0) | 0.0295 | 34,736 (20.5) | 52,860 (20.5) | 0.0004 |
|  | Neurological conditions | 6,003 (3.5) | 9,801 (3.8) | 0.0130 | 6,221 (3.7) | 9,521 (3.7) | 0.0013 |
|  | Obesity | 44,183 (26.1) | 68,847 (26.6) | 0.0118 | 44,742 (26.4) | 68,124 (26.4) | 0.0008 |
|  | Primary immunodeficiencies | 15,989 (9.5) | 25,055 (9.7) | 0.0083 | 16,167 (9.5) | 24,678 (9.6) | 0.0013 |
|  | Pregnancy<sup>b</sup> | 182 (0.1) | 357 (0.1) | 0.0087 | 215 (0.1) | 327 (0.1) | <0.0001 |
|  | Physical inactivity | 265 (0.2) | 385 (0.1) | 0.0020 | 259 (0.2) | 392 (0.2) | 0.0002 |
|  | Smoking<sup>c</sup> | 34,740 (20.5) | 54,411 (21.1) | 0.0128 | 35,290 (20.8) | 53,733 (20.8) | 0.0007 |
|  | Solid organ or hematopoietic stem cell transplant | 5,078 (3.0) | 8,679 (3.4) | 0.0203 | 5,435 (3.2) | 8,307 (3.2) | 0.0009 |
|  | Tuberculosis | 160 (0.1) | 228 (0.1) | 0.0021 | 154 (0.1) | 233 (0.1) | 0.0001 |
|  | Use of immunosuppressants | 63,796 (37.7) | 98,130 (38.0) | 0.0053 | 64,308 (37.9) | 97,664 (37.9) | 0.0010 |

IQR, interquartile range; SD, standard deviation; SMD, standardized mean difference
<sup>a</sup>except for asthma; <sup>b</sup>Includes recent pregnancy ; <sup>c</sup>Includes current and former smoker

**Supplementary Table 6.**
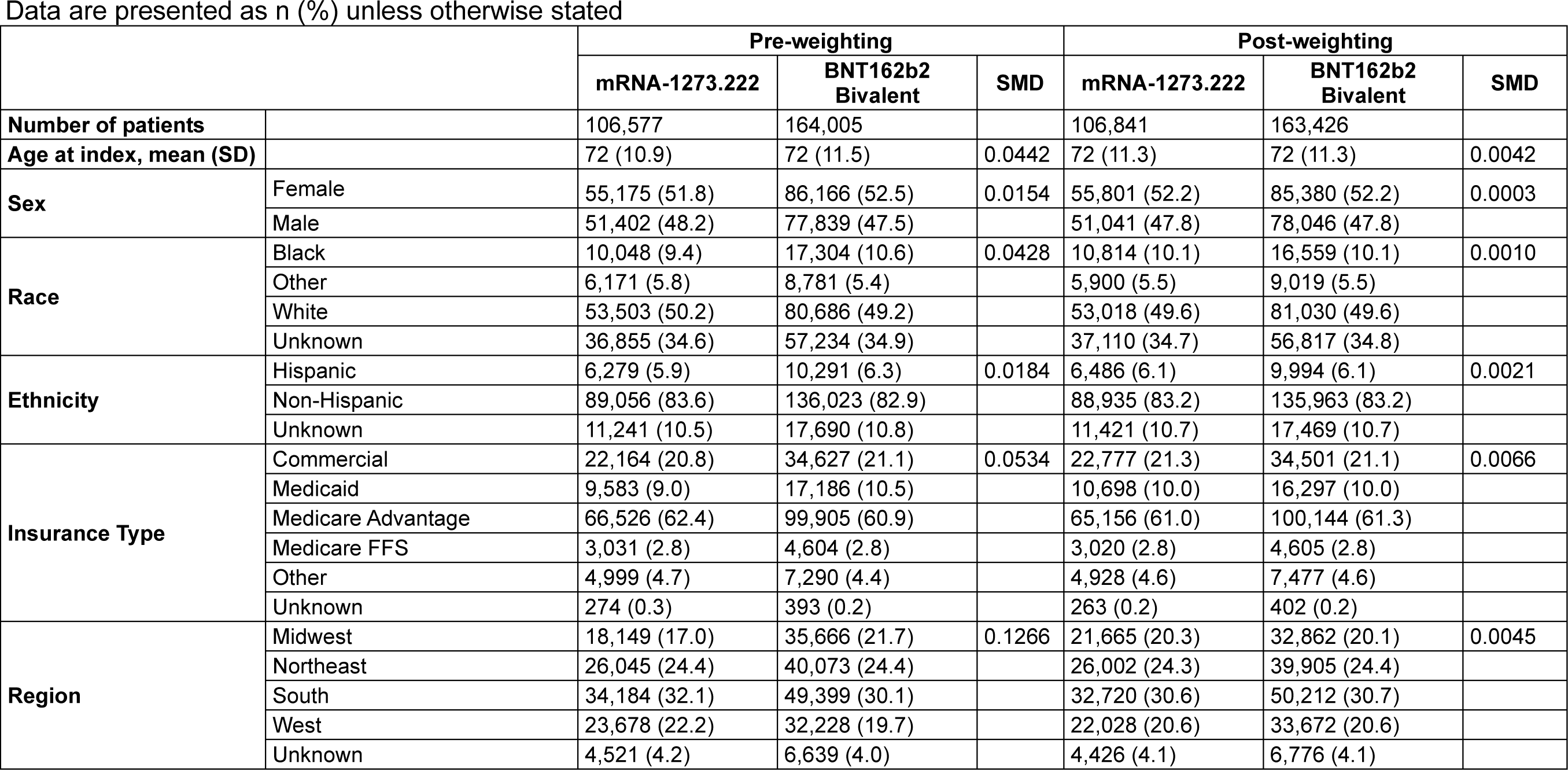
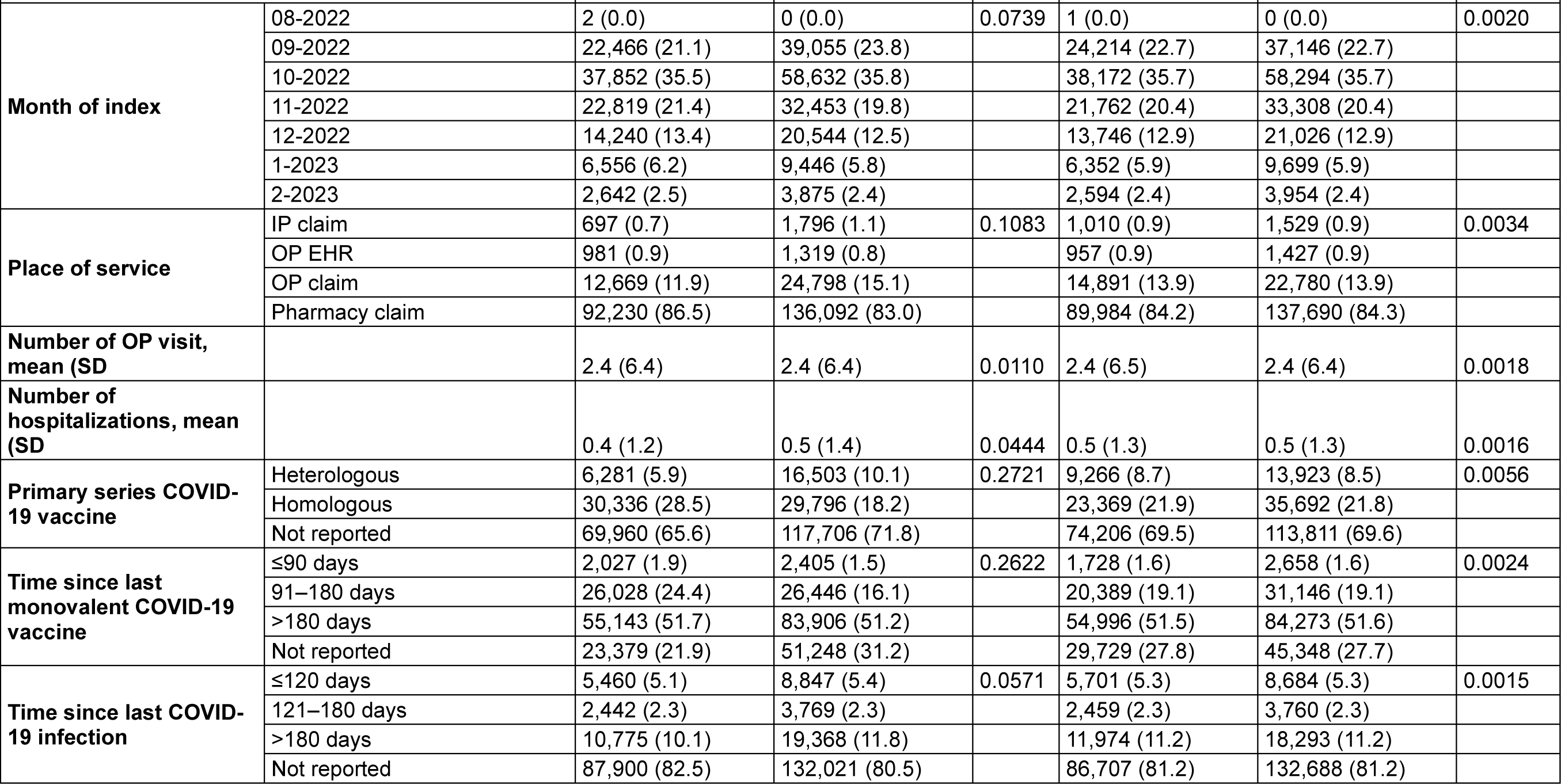
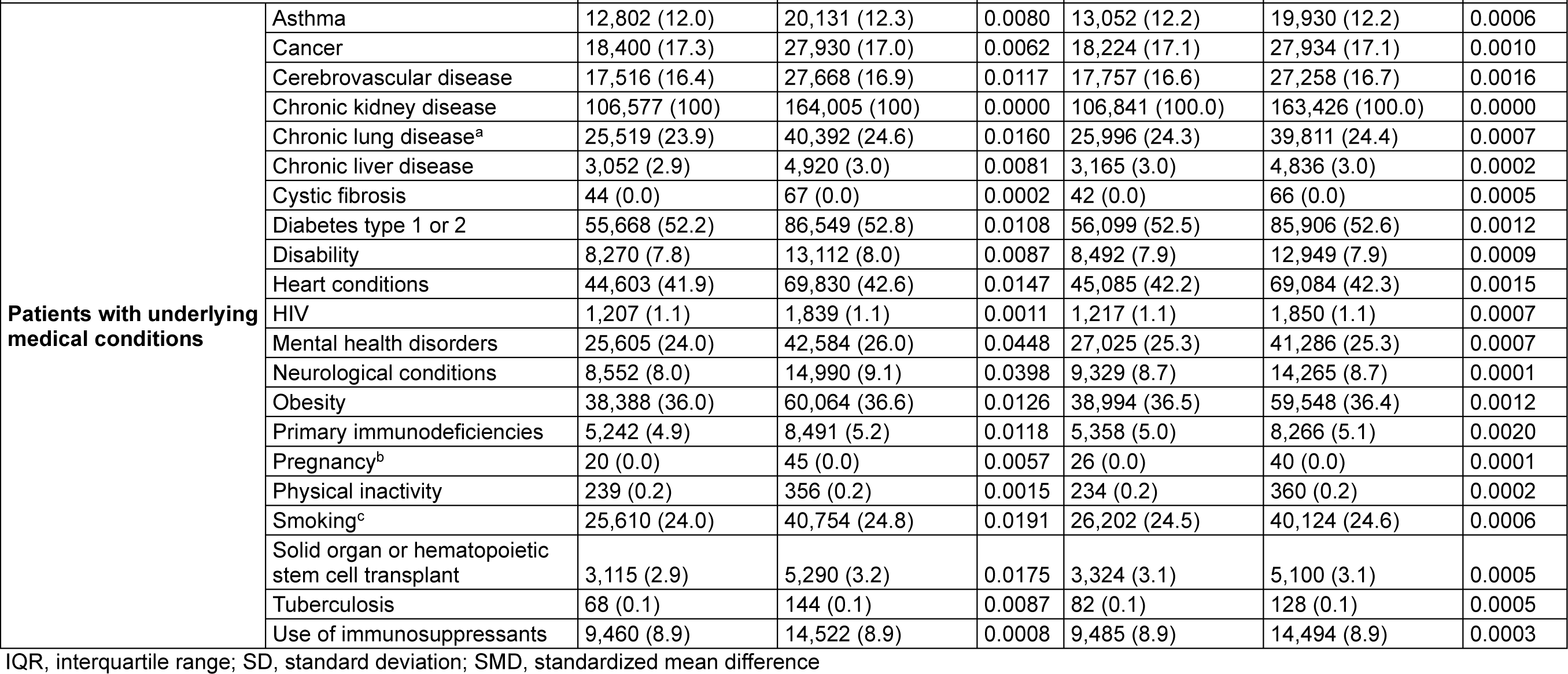
Baseline characteristics of the CKD Cohort.

Data are presented as n (%) unless otherwise stated
|  |  | Pre-weighting |  |  | Post-weighting |  |  |
| --- | --- | --- | --- | --- | --- | --- | --- |
|  |  | mRNA-1273.222 | BNT162b2 Bivalent | SMD | mRNA-1273.222 | BNT162b2 Bivalent | SMD |
| **Number of patients** |  | 106,577 | 164,005 |  | 106,841 | 163,426 |  |
| **Age at index, mean (SD)** |  | 72 (10.9) | 72 (11.5) | 0.0442 | 72 (11.3) | 72 (11.3) | 0.0042 |
| **Sex** | Female | 55,175 (51.8) | 86,166 (52.5) | 0.0154 | 55,801 (52.2) | 85,380 (52.2) | 0.0003 |
|  | Male | 51,402 (48.2) | 77,839 (47.5) |  | 51,041 (47.8) | 78,046 (47.8) |  |
| **Race** | Black | 10,048 (9.4) | 17,304 (10.6) | 0.0428 | 10,814 (10.1) | 16,559 (10.1) | 0.0010 |
|  | Other | 6,171 (5.8) | 8,781 (5.4) |  | 5,900 (5.5) | 9,019 (5.5) |  |
|  | White | 53,503 (50.2) | 80,686 (49.2) |  | 53,018 (49.6) | 81,030 (49.6) |  |
|  | Unknown | 36,855 (34.6) | 57,234 (34.9) |  | 37,110 (34.7) | 56,817 (34.8) |  |
| **Ethnicity** | Hispanic | 6,279 (5.9) | 10,291 (6.3) | 0.0184 | 6,486 (6.1) | 9,994 (6.1) | 0.0021 |
|  | Non-Hispanic | 89,056 (83.6) | 136,023 (82.9) |  | 88,935 (83.2) | 135,963 (83.2) |  |
|  | Unknown | 11,241 (10.5) | 17,690 (10.8) |  | 11,421 (10.7) | 17,469 (10.7) |  |
| **Insurance Type** | Commercial | 22,164 (20.8) | 34,627 (21.1) | 0.0534 | 22,777 (21.3) | 34,501 (21.1) | 0.0066 |
|  | Medicaid | 9,583 (9.0) | 17,186 (10.5) |  | 10,698 (10.0) | 16,297 (10.0) |  |
|  | Medicare Advantage | 66,526 (62.4) | 99,905 (60.9) |  | 65,156 (61.0) | 100,144 (61.3) |  |
|  | Medicare FFS | 3,031 (2.8) | 4,604 (2.8) |  | 3,020 (2.8) | 4,605 (2.8) |  |
|  | Other | 4,999 (4.7) | 7,290 (4.4) |  | 4,928 (4.6) | 7,477 (4.6) |  |
|  | Unknown | 274 (0.3) | 393 (0.2) |  | 263 (0.2) | 402 (0.2) |  |
| **Region** | Midwest | 18,149 (17.0) | 35,666 (21.7) | 0.1266 | 21,665 (20.3) | 32,862 (20.1) | 0.0045 |
|  | Northeast | 26,045 (24.4) | 40,073 (24.4) |  | 26,002 (24.3) | 39,905 (24.4) |  |
|  | South | 34,184 (32.1) | 49,399 (30.1) |  | 32,720 (30.6) | 50,212 (30.7) |  |
|  | West | 23,678 (22.2) | 32,228 (19.7) |  | 22,028 (20.6) | 33,672 (20.6) |  |
|  | Unknown | 4,521 (4.2) | 6,639 (4.0) |  | 4,426 (4.1) | 6,776 (4.1) |  |
|  |  | mRNA-1273.222 | BNT162b2 Bivalent | SMD | mRNA-1273.222 | BNT162b2 Bivalent | SMD |
| Month of index | 08-2022 | 2 (0.0) | 0 (0.0) | 0.0739 | 1 (0.0) | 0 (0.0) | 0.0020 |
|  | 09-2022 | 22,466 (21.1) | 39,055 (23.8) |  | 24,214 (22.7) | 37,146 (22.7) |  |
|  | 10-2022 | 37,852 (35.5) | 58,632 (35.8) |  | 38,172 (35.7) | 58,294 (35.7) |  |
|  | 11-2022 | 22,819 (21.4) | 32,453 (19.8) |  | 21,762 (20.4) | 33,308 (20.4) |  |
|  | 12-2022 | 14,240 (13.4) | 20,544 (12.5) |  | 13,746 (12.9) | 21,026 (12.9) |  |
|  | 1-2023 | 6,556 (6.2) | 9,446 (5.8) |  | 6,352 (5.9) | 9,699 (5.9) |  |
|  | 2-2023 | 2,642 (2.5) | 3,875 (2.4) |  | 2,594 (2.4) | 3,954 (2.4) |  |
| Place of service | IP claim | 697 (0.7) | 1,796 (1.1) | 0.1083 | 1,010 (0.9) | 1,529 (0.9) | 0.0034 |
|  | OP EHR | 981 (0.9) | 1,319 (0.8) |  | 957 (0.9) | 1,427 (0.9) |  |
|  | OP claim | 12,669 (11.9) | 24,798 (15.1) |  | 14,891 (13.9) | 22,780 (13.9) |  |
|  | Pharmacy claim | 92,230 (86.5) | 136,092 (83.0) |  | 89,984 (84.2) | 137,690 (84.3) |  |
| Number of OP visit, mean (SD) |  | 2.4 (6.4) | 2.4 (6.4) | 0.0110 | 2.4 (6.5) | 2.4 (6.4) | 0.0018 |
| Number of hospitalizations, mean (SD) |  | 0.4 (1.2) | 0.5 (1.4) | 0.0444 | 0.5 (1.3) | 0.5 (1.3) | 0.0016 |
| Primary series COVID-19 vaccine | Heterologous | 6,281 (5.9) | 16,503 (10.1) | 0.2721 | 9,266 (8.7) | 13,923 (8.5) | 0.0056 |
|  | Homologous | 30,336 (28.5) | 29,796 (18.2) |  | 23,369 (21.9) | 35,692 (21.8) |  |
|  | Not reported | 69,960 (65.6) | 117,706 (71.8) |  | 74,206 (69.5) | 113,811 (69.6) |  |
| Time since last monovalent COVID-19 vaccine | ≤90 days | 2,027 (1.9) | 2,405 (1.5) | 0.2622 | 1,728 (1.6) | 2,658 (1.6) | 0.0024 |
|  | 91–180 days | 26,028 (24.4) | 26,446 (16.1) |  | 20,389 (19.1) | 31,146 (19.1) |  |
|  | >180 days | 55,143 (51.7) | 83,906 (51.2) |  | 54,996 (51.5) | 84,273 (51.6) |  |
|  | Not reported | 23,379 (21.9) | 51,248 (31.2) |  | 29,729 (27.8) | 45,348 (27.7) |  |
| Time since last COVID-19 infection | ≤120 days | 5,460 (5.1) | 8,847 (5.4) | 0.0571 | 5,701 (5.3) | 8,684 (5.3) | 0.0015 |
|  | 121–180 days | 2,442 (2.3) | 3,769 (2.3) |  | 2,459 (2.3) | 3,760 (2.3) |  |
|  | >180 days | 10,775 (10.1) | 19,368 (11.8) |  | 11,974 (11.2) | 18,293 (11.2) |  |
|  | Not reported | 87,900 (82.5) | 132,021 (80.5) |  | 86,707 (81.2) | 132,688 (81.2) |  |
|  |  | mRNA-1273.222 | BNT162b2 Bivalent | SMD | mRNA-1273.222 | BNT162b2 Bivalent | SMD |
| **Patients with underlying medical conditions** | Asthma | 12,802 (12.0) | 20,131 (12.3) | 0.0080 | 13,052 (12.2) | 19,930 (12.2) | 0.0006 |
|  | Cancer | 18,400 (17.3) | 27,930 (17.0) | 0.0062 | 18,224 (17.1) | 27,934 (17.1) | 0.0010 |
|  | Cerebrovascular disease | 17,516 (16.4) | 27,668 (16.9) | 0.0117 | 17,757 (16.6) | 27,258 (16.7) | 0.0016 |
|  | Chronic kidney disease | 106,577 (100) | 164,005 (100) | 0.0000 | 106,841 (100.0) | 163,426 (100.0) | 0.0000 |
|  | Chronic lung disease<sup>a</sup> | 25,519 (23.9) | 40,392 (24.6) | 0.0160 | 25,996 (24.3) | 39,811 (24.4) | 0.0007 |
|  | Chronic liver disease | 3,052 (2.9) | 4,920 (3.0) | 0.0081 | 3,165 (3.0) | 4,836 (3.0) | 0.0002 |
|  | Cystic fibrosis | 44 (0.0) | 67 (0.0) | 0.0002 | 42 (0.0) | 66 (0.0) | 0.0005 |
|  | Diabetes type 1 or 2 | 55,668 (52.2) | 86,549 (52.8) | 0.0108 | 56,099 (52.5) | 85,906 (52.6) | 0.0012 |
|  | Disability | 8,270 (7.8) | 13,112 (8.0) | 0.0087 | 8,492 (7.9) | 12,949 (7.9) | 0.0009 |
|  | Heart conditions | 44,603 (41.9) | 69,830 (42.6) | 0.0147 | 45,085 (42.2) | 69,084 (42.3) | 0.0015 |
|  | HIV | 1,207 (1.1) | 1,839 (1.1) | 0.0011 | 1,217 (1.1) | 1,850 (1.1) | 0.0007 |
|  | Mental health disorders | 25,605 (24.0) | 42,584 (26.0) | 0.0448 | 27,025 (25.3) | 41,286 (25.3) | 0.0007 |
|  | Neurological conditions | 8,552 (8.0) | 14,990 (9.1) | 0.0398 | 9,329 (8.7) | 14,265 (8.7) | 0.0001 |
|  | Obesity | 38,388 (36.0) | 60,064 (36.6) | 0.0126 | 38,994 (36.5) | 59,548 (36.4) | 0.0012 |
|  | Primary immunodeficiencies | 5,242 (4.9) | 8,491 (5.2) | 0.0118 | 5,358 (5.0) | 8,266 (5.1) | 0.0020 |
|  | Pregnancy<sup>b</sup> | 20 (0.0) | 45 (0.0) | 0.0057 | 26 (0.0) | 40 (0.0) | 0.0001 |
|  | Physical inactivity | 239 (0.2) | 356 (0.2) | 0.0015 | 234 (0.2) | 360 (0.2) | 0.0002 |
|  | Smoking<sup>c</sup> | 25,610 (24.0) | 40,754 (24.8) | 0.0191 | 26,202 (24.5) | 40,124 (24.6) | 0.0006 |
|  | Solid organ or hematopoietic stem cell transplant | 3,115 (2.9) | 5,290 (3.2) | 0.0175 | 3,324 (3.1) | 5,100 (3.1) | 0.0005 |
|  | Tuberculosis | 68 (0.1) | 144 (0.1) | 0.0087 | 82 (0.1) | 128 (0.1) | 0.0005 |
|  | Use of immunosuppressants | 9,460 (8.9) | 14,522 (8.9) | 0.0008 | 9,485 (8.9) | 14,494 (8.9) | 0.0003 |

IQR, interquartile range; SD, standard deviation; SMD, standardized mean difference

**Supplementary Table 7.** Baseline characteristics of individuals included in the open claims sensitivity analysis.

Data are presented as n (%) unless otherwise stated
|  |  | Pre-weighting |  |  | Post-weighting |  |  |
| --- | --- | --- | --- | --- | --- | --- | --- |
|  |  | mRNA-1273.222 | BNT162b2 Bivalent | SMD | mRNA-1273.222 | BNT162b2 Bivalent | SMD |
| **Number of patients** |  | 1,960,185 | 2,969,597 |  | 1,964,137 | 2,961,234 |  |
| **Age at index, mean (SD)** |  | 66 (14.9) | 64 (16.0) | 0.1097 | 65 (15.7) | 65 (15.6) | 0.0030 |
| **Sex** | Female | 1,101,985 (56.2) | 1,700,044 (57.2) | 0.0208 | 1,116,624 (56.9) | 1,683,810 (56.9) | 0.0002 |
|  | Male | 858,200 (43.8) | 1,269,553 (42.8) |  | 847,513 (43.1) | 1,277,425 (43.1) |  |
| **Race** | Black | 120,008 (6.1) | 186,674 (6.3) | 0.0286 | 121,625 (6.2) | 184,059 (6.2) | 0.0014 |
|  | Other | 85,321 (4.4) | 128,131 (4.3) |  | 85,509 (4.4) | 128,621 (4.3) |  |
|  | White | 1,068,365 (54.5) | 1,578,195 (53.1) |  | 1,054,386 (53.7) | 1,589,130 (53.7) |  |
|  | Unknown | 686,491 (35.0) | 1,076,597 (36.3) |  | 702,616 (35.8) | 1,059,424 (35.8) |  |
| **Ethnicity** | Hispanic | 77,991 (4.0) | 128,833 (4.3) | 0.0257 | 82,148 (4.2) | 124,319 (4.2) | 0.0011 |
|  | Non-Hispanic | 1,649,332 (84.1) | 2,471,516 (83.2) |  | 1,642,149 (83.6) | 2,475,085 (83.6) |  |
|  | Unknown | 232,848 (11.9) | 369,221 (12.4) |  | 239,840 (12.2) | 361,831 (12.2) |  |
| **Insurance Type** | Commercial | 552,012 (28.2) | 897,051 (30.2) | 0.0716 | 584,112 (29.7) | 875,209 (29.6) | 0.0044 |
|  | Medicaid | 101,756 (5.2) | 183,703 (6.2) |  | 114,435 (5.8) | 172,520 (5.8) |  |
|  | Medicare Advantage | 350,121 (17.9) | 512,954 (17.3) |  | 340,184 (17.3) | 516,148 (17.4) |  |
|  | Medicare FFS | 16,044 (0.8) | 24,479 (0.8) |  | 16,126 (0.8) | 24,296 (0.8) |  |
|  | Other | 59,136 (3.0) | 95,295 (3.2) |  | 62,075 (3.2) | 93,250 (3.1) |  |
|  | Unknown | 881,116 (45.0) | 1,256,115 (42.3) |  | 847,205 (43.1) | 1,279,811 (43.2) |  |
| **Region** | Midwest | 387,534 (19.8) | 714,791 (24.1) | 0.1365 | 445,123 (22.7) | 667,829 (22.6) | 0.0027 |
|  | Northeast | 382,663 (19.5) | 601,138 (20.2) |  | 392,031 (20.0) | 592,049 (20.0) |  |
|  | South | 732,186 (37.4) | 939,965 (31.7) |  | 660,096 (33.6) | 996,832 (33.7) |  |
|  | West | 390,534 (19.9) | 616,754 (20.8) |  | 401,658 (20.4) | 605,959 (20.5) |  |
|  | Unknown | 67,268 (3.4) | 96,949 (3.3) |  | 65,228 (3.3) | 98,567 (3.3) |  |
|  |  | mRNA-1273.222 | BNT162b2 Bivalent | SMD | mRNA-1273.222 | BNT162b2 Bivalent | SMD |
| Month of index | 08-2022 | 26 (0.0) | 51 (0.0) | 0.0961 | 33 (0.0) | 47 (0.0) | 0.0015 |
|  | 09-2022 | 422,860 (21.6) | 741,749 (25.0) |  | 464,878 (23.7) | 701,215 (23.7) |  |
|  | 10-2022 | 686,911 (35.0) | 1,060,697 (35.7) |  | 695,856 (35.4) | 1,049,508 (35.4) |  |
|  | 11-2022 | 420,241 (21.4) | 579,871 (19.5) |  | 398,247 (20.3) | 599,930 (20.3) |  |
|  | 12-2022 | 261,175 (13.3) | 356,695 (12.0) |  | 246,321 (12.5) | 370,893 (12.5) |  |
|  | 1-2023 | 122,048 (6.2) | 164,472 (5.5) |  | 113,856 (5.8) | 171,758 (5.8) |  |
|  | 2-2023 | 46,924 (2.4) | 66,062 (2.2) |  | 44,946 (2.3) | 67,883 (2.3) |  |
| Place of service | IP claim | 5,716 (0.3) | 11,248 (0.4) | 0.0702 | 6,834 (0.3) | 10,281 (0.3) | 0.0021 |
|  | OP EHR | 47,934 (2.4) | 63,041 (2.1) |  | 45,604 (2.3) | 67,788 (2.3) |  |
|  | OP claim | 211,926 (10.8) | 384,009 (12.9) |  | 238,612 (12.1) | 360,295 (12.2) |  |
|  | Pharmacy claim | 1,694,609 (86.5) | 2,511,299 (84.6) |  | 1,673,087 (85.2) | 2,522,872 (85.2) |  |
| Number of OP visits, mean (SD) |  | 2.4 (6.2) | 2.3 (6.0) | 0.0137 | 2.4 (6.2) | 2.4 (6.1) | 0.0012 |
| Number of hospitalizations, mean (SD) |  | 0.2 (0.7) | 0.2 (0.8) | 0.0104 | 0.2 (0.8) | 0.2 (0.8) | 0.0004 |
| Primary series COVID-19 vaccine | Heterologous | 128,691 (6.6) | 302,676 (10.2) | 0.2330 | 176,277 (9.0) | 261,903 (8.8) | 0.0046 |
|  | Homologous | 563,137 (28.7) | 585,351 (19.7) |  | 450,126 (22.9) | 679,068 (22.9) |  |
|  | Not reported | 1,268,357 (64.7) | 2,081,570 (70.1) |  | 1,337,733 (68.1) | 2,020,264 (68.2) |  |
| Time since last monovalent COVID-19 vaccine | ≤90 days | 29,193 (1.5) | 37,634 (1.3) | 0.1523 | 26,369 (1.3) | 39,918 (1.3) | 0.0011 |
|  | 91–180 days | 377,459 (19.3) | 430,909 (14.5) |  | 317,273 (16.2) | 478,643 (16.2) |  |
|  | >180 days | 1,083,774 (55.3) | 1,642,034 (55.3) |  | 1,087,793 (55.4) | 1,640,169 (55.4) |  |
|  | Not reported | 469,759 (24.0) | 859,020 (28.9) |  | 532,703 (27.1) | 802,504 (27.1) |  |
| Time since last COVID-19 infection | ≤120 days | 70,375 (3.6) | 111,823 (3.8) | 0.0339 | 73,033 (3.7) | 109,916 (3.7) | 0.0009 |
|  | 121–180 days | 35,365 (1.8) | 55,751 (1.9) |  | 36,615 (1.9) | 54,994 (1.9) |  |
|  | >180 days | 143,640 (7.3) | 242,213 (8.2) |  | 154,766 (7.9) | 232,819 (7.9) |  |
|  | Not reported | 1,710,805 (87.3) | 2,559,810 (86.2) |  | 1,699,724 (86.5) | 2,563,505 (86.6) |  |
|  |  | mRNA-1273.222 | BNT162b2 Bivalent | SMD | mRNA-1273.222 | BNT162b2 Bivalent | SMD |
| **Patients with underlying medical conditions** | Asthma | 239,547 (12.2) | 378,092 (12.7) | 0.0155 | 246,885 (12.6) | 371,851 (12.6) | 0.0004 |
|  | Cancer | 274,094 (14.0) | 397,098 (13.4) | 0.0178 | 266,564 (13.6) | 402,460 (13.6) | 0.0006 |
|  | Cerebrovascular disease | 182,716 (9.3) | 263,756 (8.9) | 0.0153 | 176,864 (9.0) | 267,360 (9.0) | 0.0008 |
|  | Chronic kidney disease | 311,420 (15.9) | 454,246 (15.3) | 0.0163 | 302,912 (15.4) | 458,357 (15.5) | 0.0016 |
|  | Chronic lung disease<sup>a</sup> | 279,050 (14.2) | 396,105 (13.3) | 0.0260 | 267,171 (13.6) | 403,897 (13.6) | 0.0011 |
|  | Chronic liver disease | 34,511 (1.8) | 52,909 (1.8) | 0.0016 | 34,759 (1.8) | 52,529 (1.8) | 0.0003 |
|  | Cystic fibrosis | 567 (0.0) | 871 (0.0) | 0.0002 | 575 (0.0) | 867 (0.0) | <0.0001 |
|  | Diabetes type 1 or 2 | 672,173 (34.3) | 978,019 (32.9) | 0.0287 | 654,522 (33.3) | 989,186 (33.4) | 0.0017 |
|  | Disability | 169,788 (8.7) | 285,896 (9.6) | 0.0335 | 183,394 (9.3) | 275,196 (9.3) | 0.0015 |
|  | Heart conditions | 475,711 (24.3) | 679,657 (22.9) | 0.0325 | 457,340 (23.3) | 691,483 (23.4) | 0.0016 |
|  | HIV | 14,635 (0.7) | 21,945 (0.7) | 0.0009 | 14,702 (0.7) | 22,066 (0.7) | 0.0004 |
|  | Mental health disorders | 455,446 (23.2) | 751,226 (25.3) | 0.0481 | 483,565 (24.6) | 727,306 (24.6) | 0.0014 |
|  | Neurological conditions | 80,821 (4.1) | 133,357 (4.5) | 0.0181 | 85,055 (4.3) | 128,647 (4.3) | 0.0007 |
|  | Obesity | 573,660 (29.3) | 882,196 (29.7) | 0.0097 | 580,259 (29.5) | 875,008 (29.5) | 0.0001 |
|  | Primary immunodeficiencies | 36,939 (1.9) | 55,914 (1.9) | 0.0001 | 36,724 (1.9) | 55,583 (1.9) | 0.0005 |
|  | Pregnancy<sup>b</sup> | 9,095 (0.5) | 17,848 (0.6) | 0.0188 | 10,969 (0.6) | 16,367 (0.6) | 0.0008 |
|  | Physical inactivity | 2,913 (0.1) | 4,638 (0.2) | 0.0019 | 3,071 (0.2) | 4,586 (0.2) | 0.0004 |
|  | Smoking<sup>c</sup> | 373,645 (19.1) | 566,861 (19.1) | 0.0007 | 374,189 (19.1) | 564,840 (19.1) | 0.0006 |
|  | Solid organ or hematopoietic stem cell transplant | 13,433 (0.7) | 21,637 (0.7) | 0.0052 | 13,953 (0.7) | 21,098 (0.7) | 0.0002 |
|  | Tuberculosis | 945 (0.0) | 1,504 (0.1) | 0.0011 | 972 (0.0) | 1,468 (0.0) | <0.0001 |
|  | Use of immunosuppressants | 151,757 (7.7) | 225,188 (7.6) | 0.0060 | 149,962 (7.6) | 226,206 (7.6) | 0.0001 |

IQR, interquartile range; SD, standard deviation; SMD, standardized mean difference
<sup>a</sup>except for asthma; <sup>b</sup>Includes recent pregnancy ; <sup>c</sup>Includes current and former smoker

**Supplementary Table 8.** Baseline characteristics of individuals included in the sensitivity analysis using an end date of February 28, 2023.

Data are presented as n (%) unless otherwise stated
|  |  | Pre-weighting |  |  | Post-weighting |  |  |
| --- | --- | --- | --- | --- | --- | --- | --- |
|  |  | mRNA-1273.222 | BNT162b2 Bivalent | SMD | mRNA-1273.222 | BNT162b2 Bivalent | SMD |
| **Number of patients** |  | 748,343 | 1,190,324 |  | 749,557 | 1,187,593 |  |
| **Age at index, mean (SD)** |  | 62 (15.6) | 61 (16.4) | 0.0954 | 61 (16.1) | 61 (16.1) | 0.0027 |
| **Sex** | Female | 425,872 (56.9) | 689,028 (57.9) | 0.0198 | 431,148 (57.5) | 683,166 (57.5) | 0.0001 |
|  | Male | 322,471 (43.1) | 501,296 (42.1) |  | 318,409 (42.5) | 504,427 (42.5) |  |
| **Race** | Black | 46,949 (6.3) | 81,183 (6.8) | 0.0307 | 49,290 (6.6) | 78,452 (6.6) | 0.0012 |
|  | Other | 39,060 (5.2) | 59,829 (5.0) |  | 38,334 (5.1) | 60,640 (5.1) |  |
|  | White | 376,176 (50.3) | 584,959 (49.1) |  | 371,598 (49.6) | 588,544 (49.6) |  |
|  | Unknown | 286,158 (38.2) | 464,353 (39.0) |  | 290,336 (38.7) | 459,958 (38.7) |  |
| **Ethnicity** | Hispanic | 37,233 (5.0) | 62,511 (5.3) | 0.0178 | 38,418 (5.1) | 61,082 (5.1) | 0.0006 |
|  | Non-Hispanic | 624,209 (83.4) | 985,057 (82.8) |  | 622,353 (83.0) | 985,808 (83.0) |  |
|  | Unknown | 86,899 (11.6) | 142,751 (12.0) |  | 88,786 (11.8) | 140,703 (11.8) |  |
| **Insurance Type** | Commercial | 317,681 (42.5) | 522,369 (43.9) | 0.0704 | 327,605 (43.7) | 516,557 (43.5) | 0.0051 |
|  | Medicaid | 85,173 (11.4) | 154,124 (12.9) |  | 92,586 (12.4) | 147,030 (12.4) |  |
|  | Medicare Advantage | 279,592 (37.4) | 410,047 (34.4) |  | 263,539 (35.2) | 419,911 (35.4) |  |
|  | Medicare FFS | 14,509 (1.9) | 21,622 (1.8) |  | 13,940 (1.9) | 22,074 (1.9) |  |
|  | Other | 49,418 (6.6) | 79,108 (6.6) |  | 49,937 (6.7) | 78,936 (6.6) |  |
|  | Unknown | 1,970 (0.3) | 3,054 (0.3) |  | 1,950 (0.3) | 3,085 (0.3) |  |
| **Region** | Midwest | 152,244 (20.3) | 284,437 (23.9) | 0.0891 | 170,570 (22.8) | 269,236 (22.7) | 0.0023 |
|  | Northeast | 192,533 (25.7) | 303,332 (25.5) |  | 191,263 (25.5) | 303,496 (25.6) |  |
|  | South | 226,796 (30.3) | 335,353 (28.2) |  | 215,942 (28.8) | 342,916 (28.9) |  |
|  | West | 149,469 (20.0) | 226,288 (19.0) |  | 145,396 (19.4) | 230,129 (19.4) |  |
|  | Unknown | 27,301 (3.6) | 40,914 (3.4) |  | 26,386 (3.5) | 41,815 (3.5) |  |
|  |  | mRNA-1273.222 | BNT162b2 Bivalent | SMD | mRNA-1273.222 | BNT162b2 Bivalent | SMD |
| Month of index | 08-2022 | 7 (0.0) | 14 (0.0) | 0.0679 | 9 (0.0) | 13 (0.0) | 0.0011 |
|  | 09-2022 | 164,696 (22.0) | 293,437 (24.7) |  | 177,085 (23.6) | 280,869 (23.7) |  |
|  | 10-2022 | 261,030 (34.9) | 414,468 (34.8) |  | 261,220 (34.8) | 413,664 (34.8) |  |
|  | 11-2022 | 160,726 (21.5) | 240,222 (20.2) |  | 155,036 (20.7) | 245,464 (20.7) |  |
|  | 12-2022 | 101,945 (13.6) | 151,691 (12.7) |  | 98,156 (13.1) | 155,425 (13.1) |  |
|  | 1-2023 | 46,381 (6.2) | 69,596 (5.8) |  | 44,752 (6.0) | 71,025 (6.0) |  |
|  | 2-2023 | 13,558 (1.8) | 20,896 (1.8) |  | 13,298 (1.8) | 21,133 (1.8) |  |
| Place of service | IP claim | 1,975 (0.3) | 4,814 (0.4) | 0.0758 | 2,626 (0.4) | 4,196 (0.4) | 0.0014 |
|  | OP EHR | 5,138 (0.7) | 7,510 (0.6) |  | 5,021 (0.7) | 7,843 (0.7) |  |
|  | OP claim | 80,689 (10.8) | 155,641 (13.1) |  | 91,670 (12.2) | 145,459 (12.2) |  |
|  | Pharmacy claim | 660,541 (88.3) | 1,022,359 (85.9) |  | 650,240 (86.7) | 1,030,096 (86.7) |  |
| Number of OP visits, mean (SD) |  | 1.8 (5.0) | 1.7 (5.0) | 0.0108 | 1.7 (5.0) | 1.7 (5.0) | 0.0008 |
| Number of hospitalizations, mean (SD) |  | 0.2 (0.8) | 0.2 (0.9) | 0.0159 | 0.2 (0.9) | 0.2 (0.9) | 0.0009 |
| Primary series COVID-19 vaccine | Heterologous | 63,925 (8.5) | 132,120 (11.1) | 0.1754 | 78,323 (10.4) | 121,679 (10.2) | 0.0066 |
|  | Homologous | 223,982 (29.9) | 269,224 (22.6) |  | 187,906 (25.1) | 298,539 (25.1) |  |
|  | Not reported | 460,436 (61.5) | 788,980 (66.3) |  | 483,328 (64.5) | 767,375 (64.6) |  |
| Time since last monovalent COVID-19 vaccine | ≤90 days | 12,254 (1.6) | 14,892 (1.3) | 0.2316 | 10,377 (1.4) | 16,532 (1.4) | 0.0015 |
|  | 91–180 days | 157,894 (21.1) | 160,558 (13.5) |  | 121,634 (16.2) | 192,265 (16.2) |  |
|  | >180 days | 437,084 (58.4) | 715,822 (60.1) |  | 446,430 (59.6) | 707,975 (59.6) |  |
|  | Not reported | 141,111 (18.9) | 299,052 (25.1) |  | 171,115 (22.8) | 270,821 (22.8) |  |
| Time since last COVID-19 infection | ≤120 days | 32,182 (4.3) | 52,073 (4.4) | 0.0299 | 32,667 (4.4) | 51,730 (4.4) | 0.0007 |
|  | 121–180 days | 17,016 (2.3) | 27,070 (2.3) |  | 17,133 (2.3) | 27,083 (2.3) |  |
|  | >180 days | 66,145 (8.8) | 115,378 (9.7) |  | 70,366 (9.4) | 111,495 (9.4) |  |
|  | Not reported | 633,000 (84.6) | 995,803 (83.7) |  | 629,391 (84.0) | 997,285 (84.0) |  |
|  |  | mRNA-1273.222 | BNT162b2 Bivalent | SMD | mRNA-1273.222 | BNT162b2 Bivalent | SMD |
| **Patients with underlying medical conditions** | Asthma | 106,345 (14.2) | 173,045 (14.5) | 0.0093 | 108,234 (14.4) | 171,378 (14.4) | 0.0003 |
|  | Cancer | 98,649 (13.2) | 149,769 (12.6) | 0.0179 | 95,634 (12.8) | 151,832 (12.8) | 0.0008 |
|  | Cerebrovascular disease | 67,396 (9.0) | 103,404 (8.7) | 0.0112 | 65,582 (8.7) | 104,310 (8.8) | 0.0012 |
|  | Chronic kidney disease | 105,270 (14.1) | 162,009 (13.6) | 0.0132 | 102,392 (13.7) | 163,070 (13.7) | 0.0021 |
|  | Chronic lung disease<sup>a</sup> | 97,671 (13.1) | 148,729 (12.5) | 0.0167 | 94,682 (12.6) | 150,514 (12.7) | 0.0013 |
|  | Chronic liver disease | 14,144 (1.9) | 22,665 (1.9) | 0.0010 | 14,195 (1.9) | 22,543 (1.9) | 0.0003 |
|  | Cystic fibrosis | 282 (0.0) | 418 (0.0) | 0.0013 | 269 (0.0) | 427 (0.0) | <0.0001 |
|  | Diabetes type 1 or 2 | 248,850 (33.3) | 383,323 (32.2) | 0.0224 | 243,278 (32.5) | 386,456 (32.5) | 0.0018 |
|  | Disability | 82,002 (11.0) | 139,255 (11.7) | 0.0234 | 86,182 (11.5) | 136,041 (11.5) | 0.0013 |
|  | Heart conditions | 167,184 (22.3) | 253,448 (21.3) | 0.0254 | 161,512 (21.5) | 256,842 (21.6) | 0.0019 |
|  | HIV | 7,206 (1.0) | 11,201 (0.9) | 0.0023 | 7,147 (1.0) | 11,299 (1.0) | 0.0002 |
|  | Mental health disorders | 199,405 (26.6) | 338,248 (28.4) | 0.0396 | 208,868 (27.9) | 330,272 (27.8) | 0.0012 |
|  | Neurological conditions | 26,484 (3.5) | 46,607 (3.9) | 0.0199 | 28,159 (3.8) | 44,808 (3.8) | 0.0009 |
|  | Obesity | 252,077 (33.7) | 405,503 (34.1) | 0.0081 | 254,116 (33.9) | 402,872 (33.9) | 0.0004 |
|  | Primary immunodeficiencies | 15,769 (2.1) | 24,736 (2.1) | 0.0020 | 15,521 (2.1) | 24,716 (2.1) | 0.0007 |
|  | Pregnancy<sup>b</sup> | 4,998 (0.7) | 9,447 (0.8) | 0.0148 | 5,656 (0.8) | 8,915 (0.8) | 0.0005 |
|  | Physical inactivity | 1,185 (0.2) | 1,920 (0.2) | 0.0007 | 1,208 (0.2) | 1,909 (0.2) | 0.0001 |
|  | Smoking<sup>c</sup> | 151,537 (20.2) | 241,868 (20.3) | 0.0017 | 151,795 (20.3) | 240,899 (20.3) | 0.0008 |
|  | Solid organ or hematopoietic stem cell transplant | 5,007 (0.7) | 8,560 (0.7) | 0.0060 | 5,214 (0.7) | 8,312 (0.7) | 0.0005 |
|  | Tuberculosis | 435 (0.1) | 710 (0.1) | 0.0006 | 440 (0.1) | 699 (0.1) | 0.0001 |
|  | Use of immunosuppressants | 63,093 (8.4) | 97,056 (8.2) | 0.0101 | 61,829 (8.2) | 97,991 (8.3) | 0.0001 |

IQR, interquartile range; SD, standard deviation; SMD, standardized mean difference
<sup>a</sup>except for asthma; <sup>b</sup>Includes recent pregnancy ; <sup>c</sup>Includes current and former smoker

## Notes

### Competing Interest Statement

H.K., D.B.E., M. B-J., E. B., and J. M. are employees of and shareholders in Moderna Inc. A.B., J.P.W-J., N.Z., I.H.W., and M.B. are employees of Veradigm, which was contracted by Moderna and received fees for data management and statistical analyses. V.H.N., C.B., and T.D., are employees of VHN Consulting, which was contracted by Moderna to help conduct this analysis.

### Author Declarations

As a noninterventional, retrospective database study using a certified Health Insurance Portability and Accountability Act-compliant deidentified research database, approval by an institutional review board was not necessary.

